# Blood methylation pattern reflects epigenetic remodelling in adipose tissue after bariatric surgery

**DOI:** 10.1101/2024.03.08.24303962

**Authors:** Luise Müller, Anne Hoffmann, Stephan H. Bernhart, Adhideb Ghosh, Jiawei Zhong, Tobias Hagemann, Wenfei Sun, Hua Dong, Falko Noé, Christian Wolfrum, Arne Dietrich, Michael Stumvoll, Lucas Massier, Matthias Blüher, Peter Kovacs, Rima Chakaroun, Maria Keller

## Abstract

**Background:** Studies on DNA methylation following bariatric surgery have primarily focused on blood cells, while it is unclear to which extend it may reflect DNA methylation profiles in specific metabolically relevant organs such as adipose tissue (AT). Here, we investigated whether adipose tissue depots specific methylation changes after bariatric surgery are mirrored in blood.

**Methods:** Using Illumina 850K EPIC technology, we analysed genome-wide DNA methylation in paired blood, subcutaneous and omental visceral AT (SAT/OVAT) samples from nine individuals with severe obesity pre- and post-surgery.

**Findings:** The numbers and effect sizes of differentially methylated regions (DMRs) post-bariatric surgery were more pronounced in AT (SAT: 12,865 DMRs from -11.5 to 10.8%; OVAT: 14,632 DMRs from -13.7 to 12.8%) than in blood (9,267 DMRs from -8.8 to 7.7%). Cross-tissue DMRs implicated immune-related genes. Among them, 49 regions could be validated with similar methylation changes in blood from independent individuals. Fourteen DMRs correlated with differentially expressed genes in AT post bariatric surgery, including downregulation of *PIK3AP1* in both SAT and OVAT. DNA methylation age acceleration was significantly higher in AT compared to blood, but remained unaffected after surgery.

**Interpretation:** Concurrent methylation pattern changes in blood and AT, particularly in immune-related genes, suggest blood DNA methylation mirrors inflammatory state of AT post-bariatric surgery.

## 1. Introduction

Individuals with obesity have an increased risk for many common diseases, including type 2 diabetes mellitus (T2DM)^1^, cardiovascular disease^2^ and several types of cancer^3^. One suggested mechanism by which obesity leads to metabolic complications is the chronic low-grade inflammation of AT^4^. Weight loss is associated with an improvement of AT inflammation and obesity-related comorbidities and is therefore one of the main goals of obesity treatment^5,6^. Bariatric surgery represents hereby the most effective option for individuals with severe obesity. It was shown that bariatric surgery improves insulin sensitivity and can thereby reverse T2DM in up to 77% of the patients in the first two years after surgery^7,8^. Furthermore, bariatric surgery reduces chronic low-grade inflammation^9,10^, the risk of several cancer types^11^, and improves cardiovascular function^12^, but the underlying molecular pathways are not completely understood.

In obesity, WAT displays detrimental metabolic alterations, hypertrophic fat cells, ectopic fat distribution and chronic low-grade inflammation, all promoting the development of T2DM^13^. It was shown that bariatric surgery improved WAT function even five years post-surgery and in spite of weight regain^14^. The downregulation of inflammatory gene expression in WAT^15^ may be one key mechanism for metabolic improvements after surgery.

Epigenetics has been proposed as a factor mediating obesity development by providing a link between environmental factors and disease mechanisms^16,17^. Distinct DNA methylation patterns were identified in the context of obesity^18,19^ and their role in mediating obesity-related comorbidities by transcriptional regulation has been suggested^20^. Longitudinal intervention studies have identified DNA methylation patterns that respond to environmental factors, and present reversible methylation marks following weight loss^21–24^. Many studies address DNA methylation changes after bariatric surgery and indicate that epigenetic regulations may mediate beneficial effect of surgery^25–28^.

The major limitation of these studies is that DNA methylation changes after weight loss were analysed in blood only. Nevertheless, a few studies suggested that obesity-related methylation marks are comparable between blood and AT^29,30^.

Moreover, differences between DNA methylation age (DNAmAge) and chronological age, called age acceleration (ageAcc), have been associated not only with BMI and obesity^31,32^, but also with changes in lifestyle and diet^33–35^ as well as surgery-induced weight loss^27,36^. However, to date, an effect of weight loss on age acceleration has not been shown for metabolically active tissues such as AT.

In this study, we conducted a comparative analysis of DNA methylation changes within individuals across three tissues: blood, SAT, and OVAT. Our aim was to identify and validate patterns in blood methylation that could signify metabolic remodelling occurring in AT after surgery. Additionally, we assessed their biological relevance by analysing corresponding mRNA transcription changes in SAT and OVAT from a separate group of individuals undergoing a two-step bariatric surgery approach. Finally, we compared the DNAmAge before and after surgery examining differences between tissue types and exploring associations with surgery-induced phenotypic improvements.

## 2. Materials and methods

### 2.1 Study cohort and design

For the initial genome-wide DNA methylation analysis, nine individuals with severe obesity, who underwent a two-step bariatric surgery, were selected from the Leipzig Obesity BioBank (LOBB). The first surgery included vertical sleeve gastrectomy and Roux-en-Y gastric bypass (RYGB), whereas the second surgery was undertaken for different reasons. All laboratory analyses of metabolic parameters, anthropometric parameter and body composition were collected prior to both surgeries as detailed before^37,38^ (Supp. Information). Excess BMI loss (EBL, %) was calculated as follows: EBL = (BMIpre - BMIpost) / (BMIpre - 25 kg/m²) * 100^39^. The time between surgeries was 3.0 ± 2.3 years. Only individuals with a stable body weight for at least three months prior the first surgery (≤ 3% fluctuations of body weight) and an EBL of at least 30% at second surgery were included (mean EBL ± SD = 55.5 ± 14.6%). All subjects gave written informed consent prior to study participation. The study was approved by the Ethics Committee of the University of Leipzig (approval no: 159-12-21052012, 017ek-2012) and performed in accordance with the Declaration of Helsinki. Detailed cohort characteristics are described in Table 1. Summarised study design is provided in Figure 1.

**Table 1.**
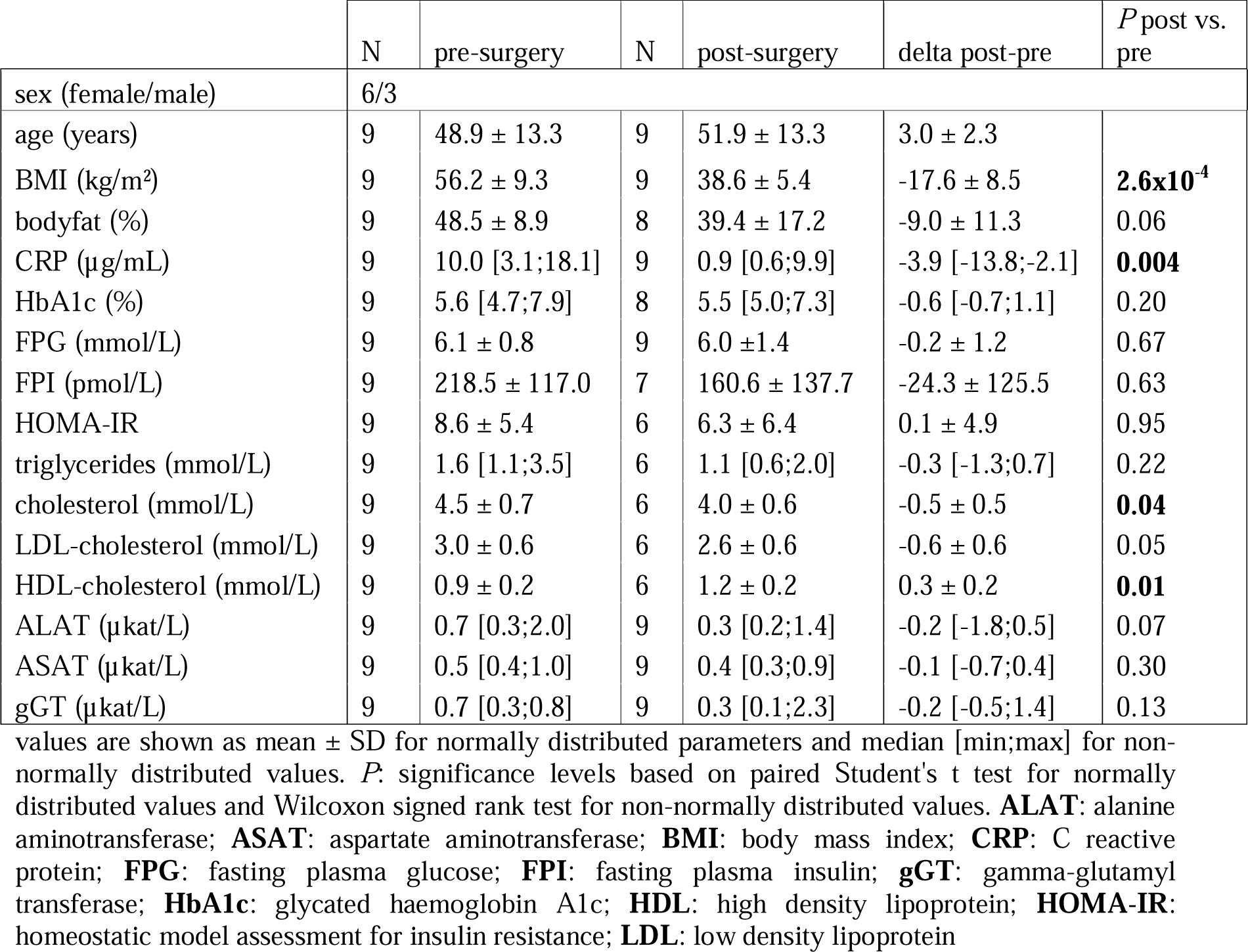
Clinical characteristics of individuals undergoing bariatric surgery.

**Figure 1.**
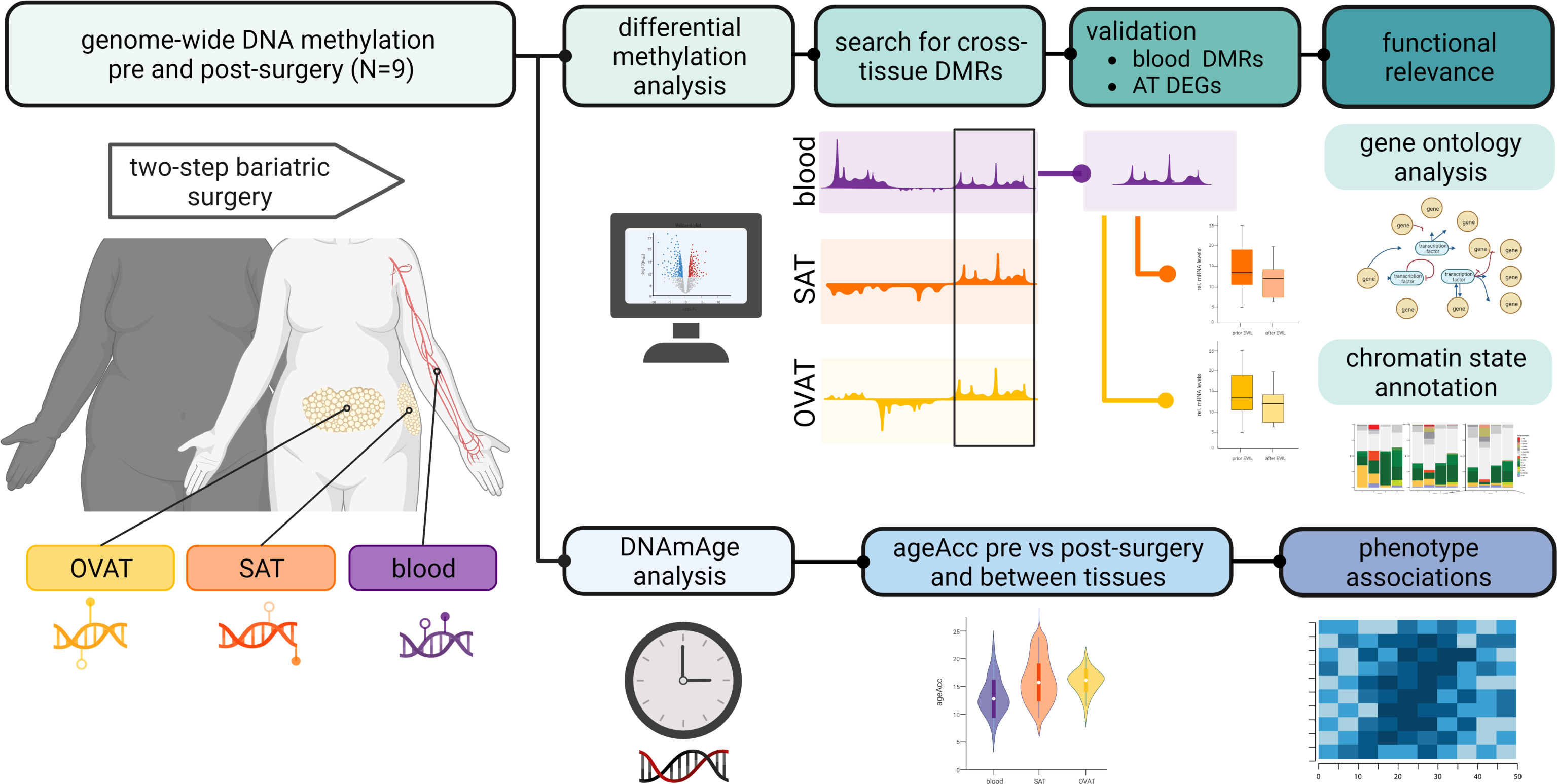
Study design. Schematic study design of initial genome-wide DNA methylation analysis in paired samples of blood, subcutaneous adipose tissue (SAT) and omental visceral adipose tissue (OVAT) from nine individuals undergoing two step bariatric surgery. Differentially methylated regions between pre and post-surgery were compared between tissues. Identified cross-tissue DMRs were validated in an independent blood sample set of individuals undergoing bariatric surgery (N = 9). Differential expression of genes (DEG) annotated to validated cross-tissue DMRs was tested in another sample set of paired SAT and OVAT samples of individuals undergoing two step bariatric surgery (N = 44). Additionally, DNA methylation age (DNAmAge) from discovery sample set was calculated by Horvath’s clock^76^. Age acceleration (ageAcc), referring to the distance between DNAmAge and chronological age, was compared between pre- and post-surgery as well as between blood, SAT and OVAT. DNAmAge pre-surgery as well as surgery induced DNAmAge changes were correlated with phenotypic changes. Figure was created with BioRender.com

For validation of blood DNA methylation marks another set of subjects from LOBB was selected. We matched nine subjects with a very close EBL after 24 months (mean EBL ± SD = 63.8 ± 5.76%, unpaired Student’s t-test of discovery vs validation *P* = 0.145; Supp. Figure S1). Blood samples were collected, and phenotypes recorded before and 24 months post-surgery. Detailed phenotypes are described in Supp. Table S1.

AT samples for the analysis of gene specific AT transcriptional regulation were selected from another independent set of individuals from the LOBB (N = 44), who also underwent a two-step bariatric surgery. Subjects with an EBL < 30%, as well as if the second surgery was more than ten years apart from the first, were excluded. The mean EBL in this cohort was not significantly different from the discovery cohort (mean EBL ± SD = 53.79 ± 16.72%, unpaired Student’s t-test of discovery vs validation *P =* 0.755, Supp. Figure S1). The timeframe between both surgeries was on average 2.09 ± 1.54 years. Detailed phenotypes are provided in Supp. Table S1.

Sample collection including nucleic acid isolation is described in Supp. Information.

### 2.3 Genome-wide DNA methylation and data processing

For genome-wide DNA methylation analysis in the discovery sample set and the validation sample set 250 ng and 500 ng genomic DNA were taken respectively for bisulfite conversion using EZ DNA Methylation Gold Kit (Zymo Research, Netherlands). After samples passed quality control, they were further amplified and hybridised on Illumina HumanMethylation850 Bead Chips (Illumina, Inc., San Diego, CA, and U.S.A). The Illumina iScan array scanner was used to quantify genome-wide DNA methylation levels at 850K CpG sites per sample on single-nucleotide resolution in collaboration with the Core Unit DNA technologies at the University of Leipzig and GenomeScan in Leiden, Netherlands.

The raw data underwent further processing including initial quality control and quantile normalisation using the *minfi* R package v1.46.0^40,41^ as well as cell type heterogeneity and batch effects testing as described in detail in Supp. Information.

### 2.4 Differential methylation analysis

The genome-wide differential methylation analysis was performed individually for each tissue, comparing postoperative samples to preoperative samples. To identify differences in methylation levels at individual CpG sites, the R package *limma* v3.56.1^42^ was utilised with default parameter settings. None of the resulting differentially methylated positions (DMPs) survived multiple testing, which is a common occurrence in intervention studies due to small methylation differences between phenotypes, even though biologically meaningful differences may exist^43,44^. Finally, DMPs with a *P*-value < 0.01 were included in the downstream analysis. Differentially methylated regions (DMRs) were obtained using *DMRcate* v2.14.0^45^. Only DMRs with more than two CpG sites and a minimum smoothed FDR < 0.05 were considered differentially methylated. Normalised M-values were utilised for both differential methylation analyses, and batch effects between the data were accounted for by incorporating patient and array effects into the linear model during the calculations. Genomic annotations of DMPs and DMRs are based on human genome build 19, considering only the highest-ranking gene.

Intersections of the DMRs with genomic annotations were summarised and visualised using the R package *annotatr* v.1.26.0^46^. Regions were annotated to CpG islands, enhancers (predicted by FANTOM5^47,48^), promoter, exons, introns, and intergenic regions.

### 2.5 Bulk RNA sequencing

To generate ribosomal RNA-depleted RNA sequencing data, we followed the SMARTseq protocol^49,50^. In summary, we enriched and reverse-transcribed the RNA using Oligo (dT) and TSO primers. For cDNA amplification, *in silico* PCR primers were utilized, and the cDNA was processed with the Nextera DNA Flex kit (Illumina, San Diego, CA, USA) using Tn5. Subsequently, all libraries were sequenced as single-end reads on a Novaseq 6000 instrument (Illumina, San Diego, CA, USA) at the Functional Genomics Center Zurich, Switzerland and analysed as described in detail in Supp. Information.

### 2.6 Differential gene expression analysis

Differential gene expression analysis was conducted using the R package *DESeq2* (v3.9)^51^. To accurately estimate effect sizes, we employed the *apeglm* fold change (FC) shrinkage estimator (v1.14.0)^52^. To account for in vitro RNA degradation, transcript integrity numbers (TINs) were estimated using the R package *RSeQC* (v4.0.0)^53^ and integrated into the linear model during differential gene expression analysis. Additionally, since paired data (pre- and post-surgery) were available for each patient, the patient ID batch was included in the linear model to minimize individual patient variance. Only DEGs with an adj. *P* < 0.05 were considered statistically significant.

### 2.7 Gene ontology (GO) enrichment and methylation age analysis

GO enrichment of DMRs was performed using the goregion function of the R package *missMethyl* v.1.34.0^54^. Significance level of enrichment results was corrected for multiple testing using FDR. The 20 most enriched GO terms were summarised using REVIGO^55^.

DNAmAge was calculated using the R package *methylclock* v.1.6.0^56^ (further details in Supp. Information).

### 2.8 ChromHMM prediction

All validated cross-tissue DMRs were aligned to chromatin segments extracted from the Epigenomic Roadmap^57^. In addition to analysing the background of all cell and tissue types, we focused on AT (AT-derived mesenchymal stem cells, mesenchymal stem cell-derived cultured adipocytes, adipocyte nuclei) and blood cells (B-cells, hematopoietic stem cells (HSC), neutrophils, natural killer cells (NK), T-cells).

### 2.9 Correlation with clinical traits

To explore potential links between *PIK3AP1* and clinical characteristics, a comprehensive analysis was conducted utilizing previously published microarray and RNA sequencing data obtained from human SAT ^15,18,58–64^. Details about the analysis and cohorts included can be found in Supp. Information and Supp. Table S2.

### 2.10 Statistical analysis

All analyses were performed using R v4.3.1 (2023-06-16)^65^. Distributions of variables were tested with Shapiro-Wilk. Comparisons of dependent groups were performed with Wilcoxon signed-rank test for non-normally distributed variables (Shapiro Wilk *P* < 0.05) and with paired Student’s t-test for normally distributed variables (Shapiro Wilk *P* ≥ 0.05). Non-normally distributed variables are given as median and range [min; max], whereas normally distributed continuous variables are given as mean ± *SD*. Statistical significance was defined for *P* < 0.05. In case of multiple testing FDR < 0.05 was considered significant. Bivariate correlation analyses were performed using Spearman’s rank correlation test for non-normally distributed variables and Pearson correlation coefficient for normally distributed variables. Linear regression analysis was calculated to predict the relationship between DNAmAge (dependent variable) and chronological age. Two-way ANOVA was calculated to test the effect of time point and tissue type on ageAcc.

### 2.11 Role of the Funders

The funders had no role in the design of the study; in the collection, analyses, or interpretation of data; in the writing of the manuscript; or in the decision to publish the results.

## 3. Results

### Improvement in clinical parameters after surgery

Individuals of the discovery sample set reduced their BMI significantly by 17.6 ± 8.5 kg/m² (*P =* 2.6 x 10^-4^) (Table 1) during a time span of 0.8 - 6.7 years after the first surgery. They lost on average 55.5 ± 14.6% of their excess BMI (EBL, %). High sensitivity CRP was significantly lower after surgery (-3.9 [-13.8; -2.1] mg/L, *P =* 0.004), as well as total cholesterol (-0.5 ± 0.5 mmol/L, *P =* 0.04). In line with this, HDL-cholesterol significantly increased (0.3 ± 0.2 mmol/L, *P =* 0.01). Although not statistically significant, we observed trends for reduced body fat (%), LDL-cholesterol (mmol/l) and ALAT (µkat/l), all *P <* 0.07.

Likewise, the individuals of the validation sample set significantly reduced their BMI by 17.7 ± 3.9 kg/m² (*P =* 7.89 x 10^-7^) 24 months after surgery (Supp. Table S1). The average EBL was 63.8 ± 5.76%. Also, the individuals from the AT expression analysis significantly reduced their BMI after surgery (-14.6 [-39; -5.2] kg/m², *P* = 7.88 x 10^-9^), with an average EBL of 53.79 ± 16.72%.

### DNA methylation changes and their genomic distribution after surgery

After comparison of DNA methylation post-surgery with pre-surgery, we did not detect DMPs after surgery reaching genome-wide significance (FDR < 0.05). However, applying |log_2_ FC| > 1 and an unadjusted *P* < 0.01 as cut offs, we detected 364 DMPs in blood (Supp. Table S3), 648 DMPs in SAT (Supp. Table S4) and 512 DMPs in OVAT (Supp. Table S5). We concentrated on DMRs defined as regions with several adjacent DMPs, which are more likely to be associated with transcriptional regulation^66^. In general, we detected 9,267 DMRs in blood with mean methylation changes from -8.8 to 7.7% (Supp. Table S6), 12,865 DMRs in SAT ranging from -11.5 to 10.8% (Supp. Table S7) and 14,632 DMRs in OVAT ranging from -13.7 to 12.8% methylation difference (Supp. Table S8) after surgery. Volcano plots for all three tissues highlighting the top 20 genes annotated to the most significant DMRs are depicted in Supp. Figure S2A-C. While in AT more DMRs were hyper- (OVAT: N = 7,914, SAT N = 6,597) than hypomethylated (OVAT: N = 6,718, SAT N = 6,268), in blood more regions were hypo- (N = 5,415) than hypermethylated (N = 3,852).

Next, we intersected DMRs with genomic annotations, which revealed similar distribution patterns for all tissues (Figure 2A). Most DMRs intersect with intronic, exonic and promoter regions. However, for all genomic features, more hypomethylated DMRs were found in blood after surgery, while in OVAT more hypermethylated DMRs are detected. In SAT, more hypermethylated DMRs are found in enhancer regions, whereas hypomethylated DMRs are more prevalent in CpG islands.

**Figure 2.**
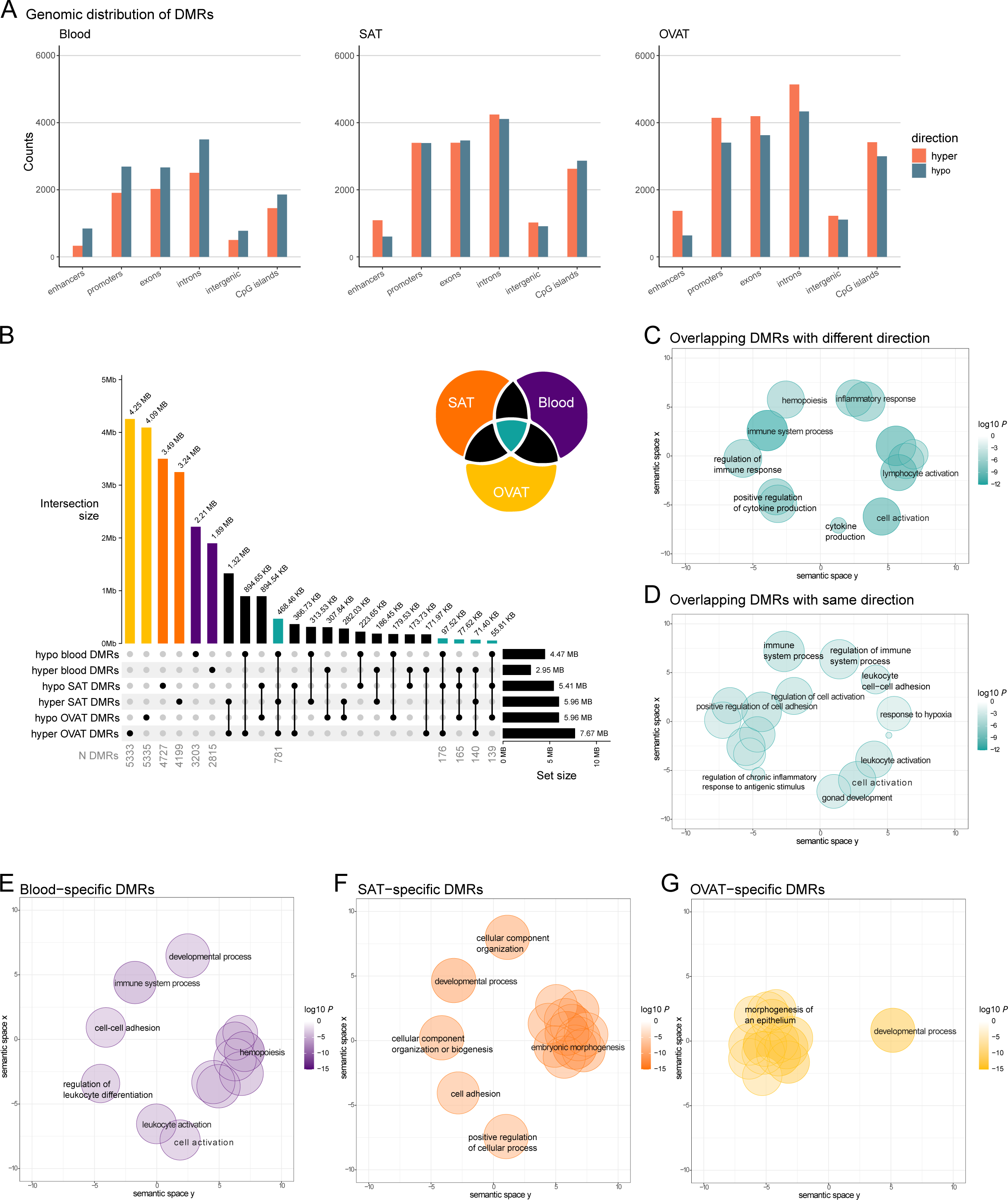
Differentially methylated regions after bariatric surgery and their genomic annotation across tissues. **A)** Genomic distribution of differentially methylated regions (DMRs) in blood, subcutaneous adipose tissue (SAT) and omental visceral adipose tissue (OVAT) after surgery. DMRs were annotated to genomic regions using annotatr. Barplot illustrates the counts of hypermethylated (depicted by red bars) and hypomethylated (depicted by blue bars) DMRs in different genomic components. DMRs covering multiple genomic components were counted in each relevant component. Enhancer data were sourced from the FANTOM database^47,48^. **B)** UpSet plot shows the intersections of DMRs across tissues separated into hypo- and hypermethylated DMRs. The top bar plot shows all intersection sizes in base pairs of overlapping DMRs for all combinations of DMR sets with an intersection size > 50 Kb. Right barplot shows the total size of DMRs for each set. Number of DMRs is displayed in grey below selected combinations. UpSet plot was generated using the R package *ComplexHeatmap*. The Venn diagram in the upper right corner illustrates the colour code for highlighted intersections. **C-G**) Results of the gene ontology (GO) enrichment analyses were summarised and visualised using REVIGO^55^. For all sets the input into REVIGO were the top 20 most significantly enriched GO terms. Scatter plots show cluster representatives after redundancy reduction in a two-dimensional space derived by applying multidimensional scaling to a matrix of the GO terms’ semantic similarities. Enriched GO terms related to biological processes of DMRs overlapping across all three tissues are visualised as scatter plots in petrol for discordant (**C**) and concordant (**D**) DMRs in blood and AT. Enriched GO terms related to biological processes of tissue-specific DMRs are shown as scatter plots in purple for blood (**E**), in orange for SAT (**F**), and in yellow for OVAT (**G**). The colour intensities of bubbles indicate the log10 enrichment *P* and the size of bubbles depicts the frequency of the GO term in the underlying GOA database, whereby bubbles of more general terms are larger and of more specialised terms are smaller.

### Overlap of DMRs between blood and adipose tissues

The upset plot (Figure 2B) visualises genomic intersections between AT- and blood methylation changes after bariatric surgery. OVAT showed the largest hypermethylated (7.67 Mb) and hypomethylated (5.96 Mb) region of DMRs, followed by SAT (hypermethylated DMRs - 5.96 Mb/ hypomethylated DMRs - 5.41 Mb). Blood DMRs were in total smaller with 4.47 Mb hypomethylated and 2.95 Mb hypermethylated regions.

The majority of DMRs were not overlapping between any of the three tissues (Figure 2B). We found 55.4% (4.25 Mb of 7.67 Mb hypermethylated DMRs) to 68.6% (4.09 Mb of 5.96 Mb hypomethylated DMRs) OVAT-specific DMRs, 54.3% (3.24 Mb from 5.96 Mb hypermethylated DMRs) to 64.5% (3.49 Mb from 5.41 Mb hypomethylated DMRs) SAT-specific DMRs, and 49.4% (2.21 Mb from 4.47 Mb hypomethylated DMRs) to 64.1% (1.89 Mb from 2.95 Mb hypermethylated DMRs) blood-specific DMRs.

We observed the highest overlap of DMRs between SAT and OVAT (1.32 Mb hypermethylated/ 894.54 Kb hypomethylated DMR intersection size). When comparing blood DMRs with AT DMRs, we found the largest overlap between hypomethylated DMRs in blood with hypermethylated DMRs in OVAT (894.65 Kb). The intersection size of DMRs overlapping in all three tissues was highest for blood and AT DMRs in different directions (468.46 Kb intersection between hypomethylated blood DMRs and hypermethylated AT DMRs, 77.62 Kb intersection between hypermethylated blood and hypomethylated AT DMRs). The intersection size of DMRs overlapping in all three tissues with the same direction was only 71.40 Kb for hypermethylated and 55.81 Kb for hypomethylated DMRs, corresponding to 2.4% of all hypermethylated blood DMRs and 1.2% of all hypomethylated blood DMRs. A detailed list of all DMRs overlapping between all three tissues (N = 1,587) can be found in Supp. Table S9.

### Gene ontology term enrichment of cross-tissue DMRs

Next, we performed gene ontology (GO) analysis for the underlying genetic annotations of regions which were targeted by changes in all three tissues (petrol-coloured bars in Figure 2B). In total 1,587 DMRs are overlapping in at least 1 bp between all three tissues regardless of their direction. The majority of intersecting DMRs were hypomethylated in blood and hypermethylated in SAT and OVAT (N = 781 DMRs), while 165 intersections were observed between hypermethylated blood DMRs and hypomethylated SAT and OVAT DMRs. The discordant DMRs between blood and both ATs were annotated to genes enriched in GO terms for immune response, lymphocyte activation, inflammatory response, and hemopoiesis, but also cytokine production and cell activation (all FDR < 0.001, Supp. Table S10, Figure 2C). From the intersecting DMRs 279 regions showed the same effect direction between all three tissues (140 hypermethylated regions, 139 hypomethylated regions). Enriched GO terms of annotated genes to those concordant DMRs were cell activation, immune system process (all FDR = 0.05, Supp. Table S10, Figure 2D). None of the enriched terms for concordant DMRs achieved an FDR < 0.05.

### Gene ontology term enrichment of tissue-specific DMRs

Additionally, we performed GO analysis for the tissue-specific DMRs. In blood 64% DMRs (5,918 from 9,267 DMRs) did not overlap with DMRs in any of the ATs. These regions were annotated to genes enriched for GO biological processes characteristic for blood, such as hemopoiesis, leukocyte differentiation and immune system process (all FDR < 0.01, Supp. Table S11, Figure 2E). In ATs around 70% of the DMRs were found to be specific for each tissue (8,926 from 12,865 SAT DMRs and 10,668 from 14,632 OVAT DMRs). Of note, for both AT depots these DMRs were annotated to genes related to developmental process and morphogenesis (all FDR < 0.001, Supp. Table S11, Figure 2F+G).

### Validated blood DMRs relate to differentially expressed genes in AT

In the genome-wide DNA methylation analysis of the validation subgroup for blood DMRs, we detected 49 DMRs overlapping with our candidate cross-tissue DMRs which showed the same effect direction as our discovery blood samples (Supp. Table S12).

To further examine the functional relevance of the validated cross-tissue DMRs we checked the mRNA expression changes of the corresponding annotated genes. From 49 validated DMRs, 14 regions were annotated to genes differentially expressed after weight loss surgery in at least one AT depot (Table 2). Ten genes in SAT and five genes in OVAT were differentially expressed after surgery. *Phosphoinositide-3-Kinase Adaptor Protein 1* (*PIK3AP1*), a gene involved in the regulation of inflammatory response (GO:0050727) was the only differentially expressed gene in both fat depots. The expression of *PIK3AP1* was significantly lower in both fat depots after surgery (log_2_ FC SAT = - 0.349, *P* adj. = 0.023; log_2_ FC OVAT = -0.769, *P* adj. = 0.0005), while the mean methylation of the DMR was increased in both fat depots after surgery (mean methylation difference in SAT = 1.59%, min smoothed FDR = 0.003; mean methylation difference in OVAT = 1.25%, min smoothed FDR = 7.78 x 10^-7^). Although we observed a lower effect size, the same DMR was also hypermethylated after surgery in blood (mean methylation difference in discovery cohort = 0.05%, min smoothed FDR = 0.002; mean methylation difference in validation cohort = 0.88%, min smoothed FDR = 0.035). The observed DMR is located in the promoter region of the *PIK3AP1* close to the transcription start site. Potential associations of *PIK3AP1* gene expression in SAT with obesity related traits were examined using previously published microarray and RNA sequencing data. Significant positive correlations were found in individual cohorts as well as in the meta-analysis of those cohorts with BMI, fat cell volume, waist to hip ratio, HOMA-IR, fasting plasma insulin and glucose, triglyceride levels and CRP levels, while significant negative correlations are found with HDL-cholesterol levels, as well as stimulated lipolysis rates in adipocytes (all *P* < 0.01, Figure 3A). Additionally, studies of Kerr et al. and Petrus et al. show consistently reduced *PIK3AP1* gene expression in SAT two and five years after bariatric surgery^15,62^ (Figure 3B) which was accompanied by reduced BMI in these subjects (Supp. Figure S3).

**Table 2.** Overview of 14 differentially expressed genes in SAT and/or OVAT annotated to validated DMRs after bariatric surgery.

|  | Overlapping region of cross-tissue DMRs |  |  | Mean methylation difference of DMR |  |  |  | DGE SAT |  |  |  | DGE OVAT |  |  |  |
| --- | --- | --- | --- | --- | --- | --- | --- | --- | --- | --- | --- | --- | --- | --- | --- |
| gene symbols | chr | start | end | Blood discovery | SAT discovery | OVAT discovery | Blood validation | log FC | SE | *P* | *P* adj. | log FC | SE | *P* | *P* adj. |
| *EML6* | 2 | 54951307 | 54951562 | -0.0012 | -0.0001 | -0.0025 | -0.0044 | -0.439 | 0.206 | 0.009 | **0.039** | -0.136 | 0.167 | 0.097 | 0.325 |
| *SLC39A14* | 8 | 22225159 | 22225395 | -0.0142 | -0.0010 | 0.0131 | -0.0152 | 0.604 | 0.216 | 0.001 | **0.009** | 0.002 | 0.107 | 0.965 | 0.984 |
| *PCBD1* | 10 | 72648646 | 72648693 | -0.0172 | 0.0011 | 0.0047 | -0.0029 | 0.265 | 0.154 | 0.050 | 0.118 | 0.469 | 0.169 | 0.000 | **0.018** |
| *LGALS3* | 14 | 55595227 | 55595969 | -0.0140 | 0.0001 | 0.0136 | -0.0110 | 0.358 | 0.156 | 0.009 | **0.039** | -0.143 | 0.127 | 0.020 | 0.160 |
| *SNRPB2* | 20 | 16710618 | 16710841 | -0.0046 | -0.0102 | 0.0040 | -0.0007 | 0.376 | 0.115 | 0.000 | **0.008** | 0.136 | 0.099 | 0.076 | 0.295 |
| *AIM2* | 1 | 159046391 | 159047177 | 0.0212 | 0.0485 | 0.0117 | 0.0264 | 0.066 | 0.199 | 0.673 | 0.760 | -1.220 | 0.331 | 0.000 | **0.001** |
| *LGALS3L* | 2 | 64681733 | 64681735 | 0.0046 | -0.0133 | -0.0115 | 0.0046 | 0.258 | 0.111 | 0.012 | **0.046** | 0.066 | 0.093 | 0.357 | 0.587 |
| *APLF* | 2 | 68694168 | 68695071 | 0.0104 | 0.0076 | 0.0061 | 0.0181 | -0.331 | 0.102 | 0.001 | **0.009** | -0.187 | 0.123 | 0.033 | 0.205 |
| *HNRNPH1* | 5 | 179050530 | 179050963 | 0.0011 | 0.0010 | -0.0131 | 0.0074 | 1.080 | 0.302 | 0.000 | **0.002** | -0.293 | 0.330 | 0.019 | 0.157 |
| *RPL7* | 8 | 74206281 | 74206317 | 0.0104 | 0.0084 | 0.0028 | 0.0042 | -1.443 | 0.037 | 0.004 | **0.026** | 0.056 | 0.120 | 0.436 | 0.652 |
| *PIK3AP1* | 10 | 98479703 | 98479991 | 0.0005 | 0.0159 | 0.0125 | 0.0088 | -0.349 | 0.131 | 0.004 | **0.023** | -0.769 | 0.184 | 0.000 | **0.001** |
| *EFEMP2* | 11 | 65640375 | 65641043 | 0.0025 | -0.0080 | -0.0066 | 0.0220 | 0.253 | 0.156 | 0.063 | 0.138 | 0.318 | 0.132 | 0.002 | **0.045** |
| *IVD* | 15 | 40697419 | 40697447 | 0.0225 | -0.0027 | -0.0181 | 0.0029 | 0.258 | 0.099 | 0.006 | **0.030** | 0.109 | 0.094 | 0.137 | 0.376 |
| *NXN* | 17 | 836474 | 837017 | 0.0281 | -0.0616 | -0.0813 | 0.0184 | 0.015 | 0.121 | 0.891 | 0.925 | 0.332 | 0.120 | 0.001 | **0.024** |

Table of genes annotated to validated DMRs differentially expressed in SAT or OVAT after bariatric surgery. Significant adjusted *P* (<0.05) of differential gene expression are marked bold. **DGE** - differential gene expression, **chr** - chromosome, **DMR** - differentially methylated region, **FC** - fold change, **OVAT** - omental visceral adipose tissue, **SAT** - subcutaneous adipose tissue.

**Figure 3.**
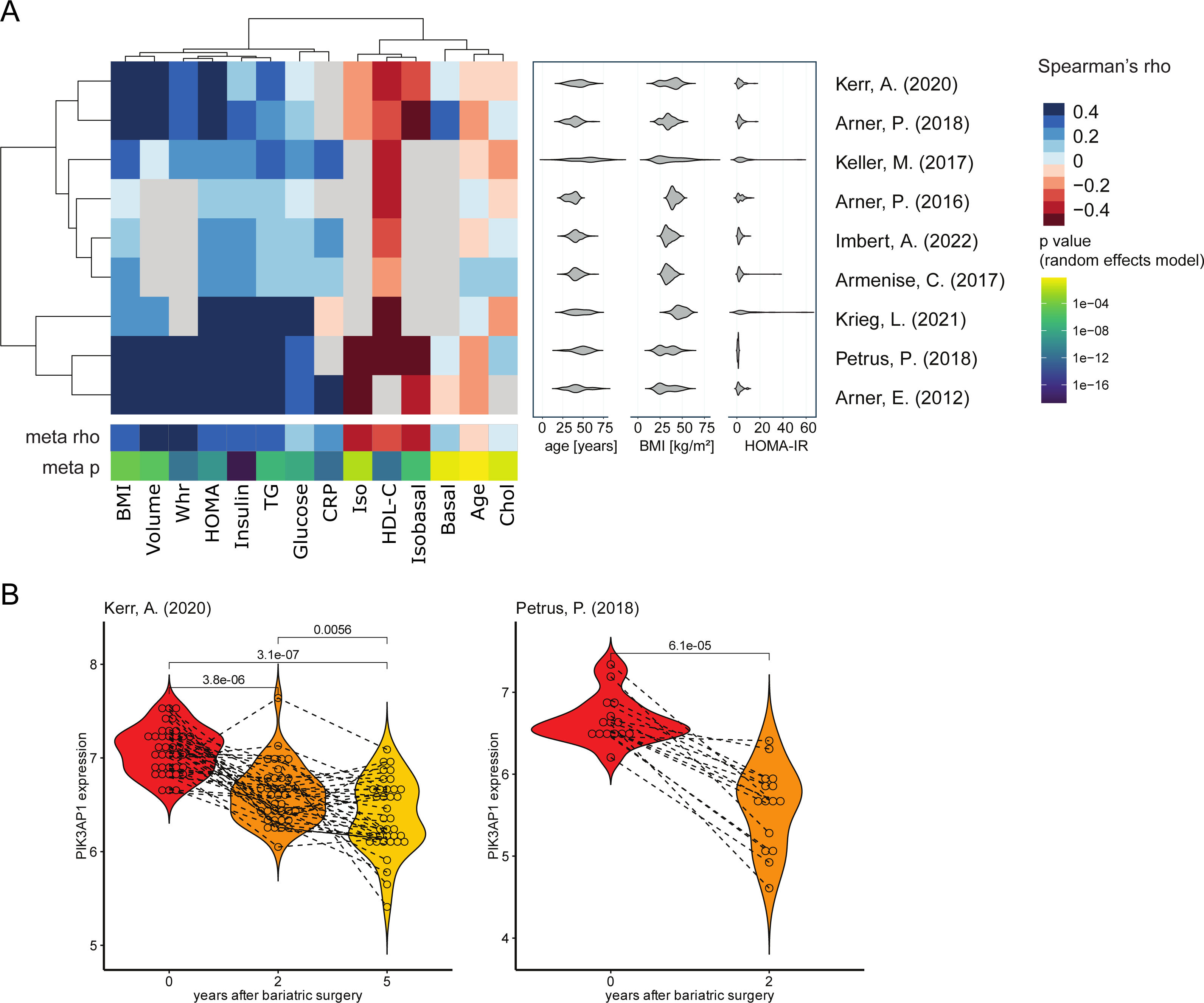
Association of *PIK3AP1* gene expression in subcutaneous adipose tissue with clinical traits and weight loss after bariatric surgery. A) Previously published microarray and RNA sequencing data from subcutaneous adipose tissue (SAT) samples were used to calculate Spearman correlation between *PIK3AP1* gene expression and the following clinical traits: body mass index (BMI), fat cell size (Volume), waist to hip ratio (WHR), homeostasis model assessment (HOMA), fasting plasma insulin (Insulin), serum triglyceride levels (TG), fasting plasma glucose (Glucose), C-reactive protein (CRP), stimulated lipolysis of isolated fat cells (Iso), high density lipoprotein cholesterol (HDL-C), ratio of stimulated to spontaneous lipolysis of fat cells (Isobasal), spontaneous lipolysis (Basal), Age and total cholesterol (Chol). The characteristics (Age, BMI and HOMA) of each cohort is shown by violin plot next to correlation matrix. Results of the meta-analysis from all cohorts for each trait (meta rho, meta p) are shown below correlation matrix. Negative correlations are shown in red and positive correlations are shown in blue. B) Violin plots show *PIK3AP1* gene expression in SAT before and after bariatric surgery in two previously published studies. Normalized and log2-transformed data were compared applying a Wilcoxon signed rank test for paired samples.

### Validated blood DMRs map in regulatory regions for gene transcription

Finally, we further proved the functional relevance of the 14 validated DMRs by chromatin state prediction in adipocytes and blood cells (Figure 4). Most validated DMRs are located predominantly in the transcriptional active regions in both adipocytes as well as blood cells, such as promoter (TssA/TssAFlnk) and enhancer regions (Enh). However, some DMRs show different chromatin states across adipocytes and blood cells. The DMR of *PIK3AP1* is predicted to be a transcriptionally active region (TssA, TssAFlnk, Enh) in blood cells, while in adipocytes this region is predicted to be less transcriptionally active (TssBiv, BivFlnk, EnhBiv, repressed PolyComb (ReprPc)). In addition, the DMR at *Absent in melanoma 2 (AIM2)* is located in most blood cells at a predicted promoter region (TssA, TssAFlnk), but in AT cells in a less transcriptional regulatory region (weak transcription (TxWk), weak repressed PolyComb (ReprPcWk), quiescent (Quies)). This goes in line with the predominant expression of both genes in blood and immune cells (Human Protein Atlas, version 23.0^67,68^). Another DMR at *Nucleoredoxin* (*NXN*) is predicted to be a transcriptional regulatory region (TSS, TssAFlnk, Enh) in adipocytes, while in blood this region is predicted to be not regulatory (ReprPcWk, Quies). Although its expression is not tissue-specific, it was found to be enriched in visceral adipose progenitor cells (Human Protein Atlas, version 23.0^69^).

**Figure 4.**
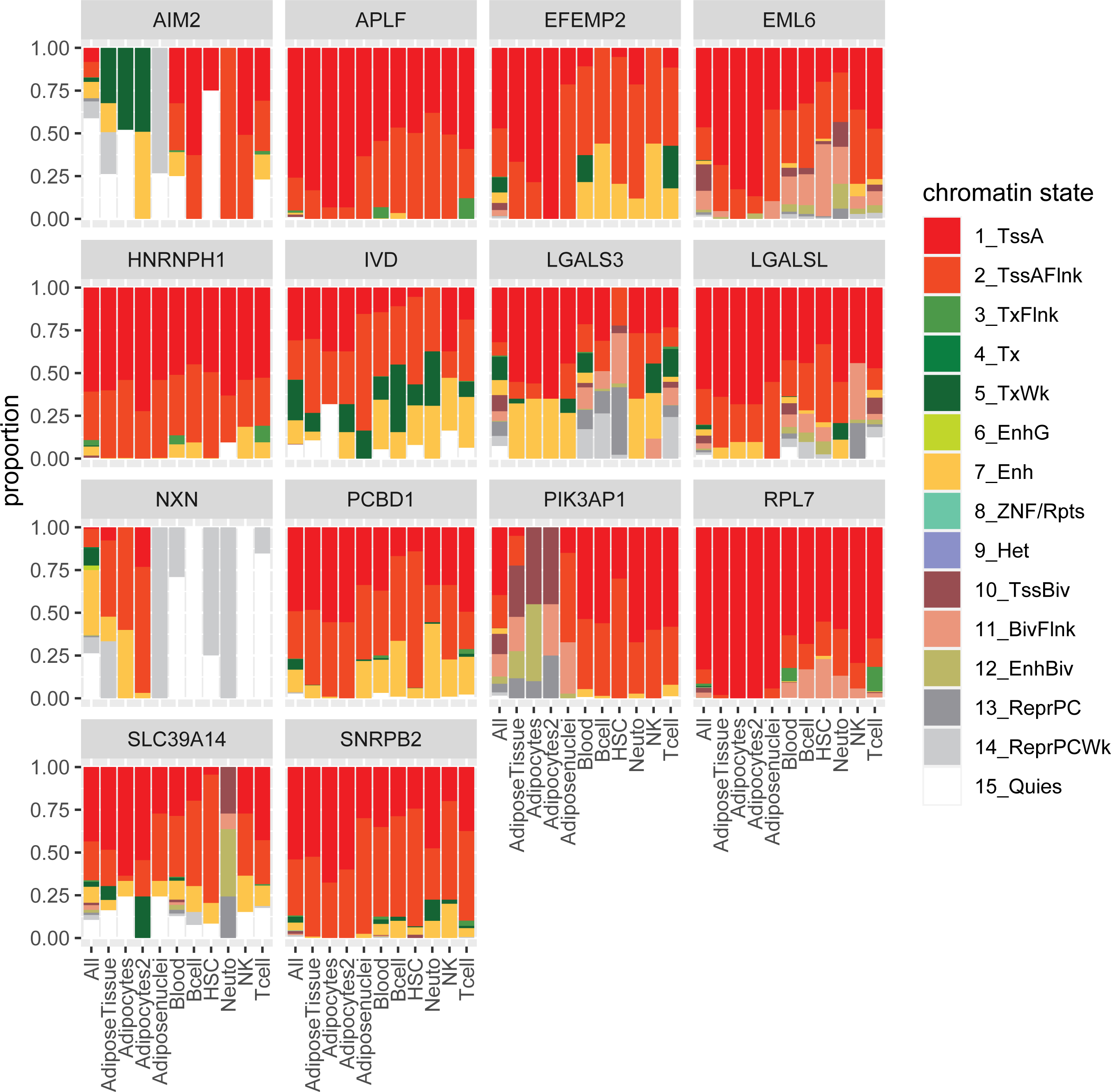
Chromatin state prediction of validated differentially methylated regions in different adipocytes and blood cells. Barplot shows the proportions of predicted chromatin states in adipocytes and blood cells of the validated differentially methylated regions (DMRs) annotated to differential expressed genes in adipose tissue. Active states (associated with expressed genes) consist of active TSS-proximal promoter states (TssA, TssAFlnk), a transcribed state at the 5’ and 3’ end of genes showing both promoter and enhancer signatures (TxFlnk), actively transcribed states (Tx, TxWk), enhancer states (Enh, EnhG), and a state associated with zinc finger protein genes (ZNF/Rpts). The inactive states consist of constitutive heterochromatin (Het), bivalent regulatory states (TssBiv, BivFlnk, EnhBiv), repressed Polycomb states (ReprPC, ReprPCWk), and a quiescent state (Quies).

### DNA methylation age acceleration is higher in adipose tissue

In all three tissues, the DNAmAge correlated significantly with the chronological age given by the date of birth (Supp. Figure S4A, all *P* < 0.001). Linear regression model between DNAmAge and chronological age fitted well for AT samples (OVAT: pre R² = 0.90, post R² = 0.97; SAT: pre R² = 0.94, post R² = 0.93), as well as blood samples (pre R² = 0.91, post R² = 0.89). There was no significant interaction of the surgery effect and the tissue type on ageAcc in our sample set using two-way repeated measures ANOVA. In line, ageAcc did not change after surgery in any of the three tissues (paired Student’s t-test, all FDR > 0.05), which might be due to the small sample size (N = 9) (Figure 5A). However, we identified a tissue-specific effect on ageAcc (Figure 5B), confirming a higher ageAcc of both AT depots compared to blood at both timepoints. Correlation analyses (Pearson) show that ageAcc in blood correlated stronger with ageAcc of each fat depot before surgery (blood vs OVAT: R = 0.97, *P* < 0.001; blood vs SAT: R = 0.89, *P* < 0.002) than after (blood vs OVAT: R = 0.6, n.s.; blood vs SAT: R = 0.68, *P* < 0.05) (Supp. Figure S4B/C). However, ageAcc of both fat depots correlated significantly (pre: R = 0.87, *P* < 0.01), which was not affected by surgery (post: R = 0.8, *P* < 0.01) (Supp. Figure S4D).

**Figure 5.**
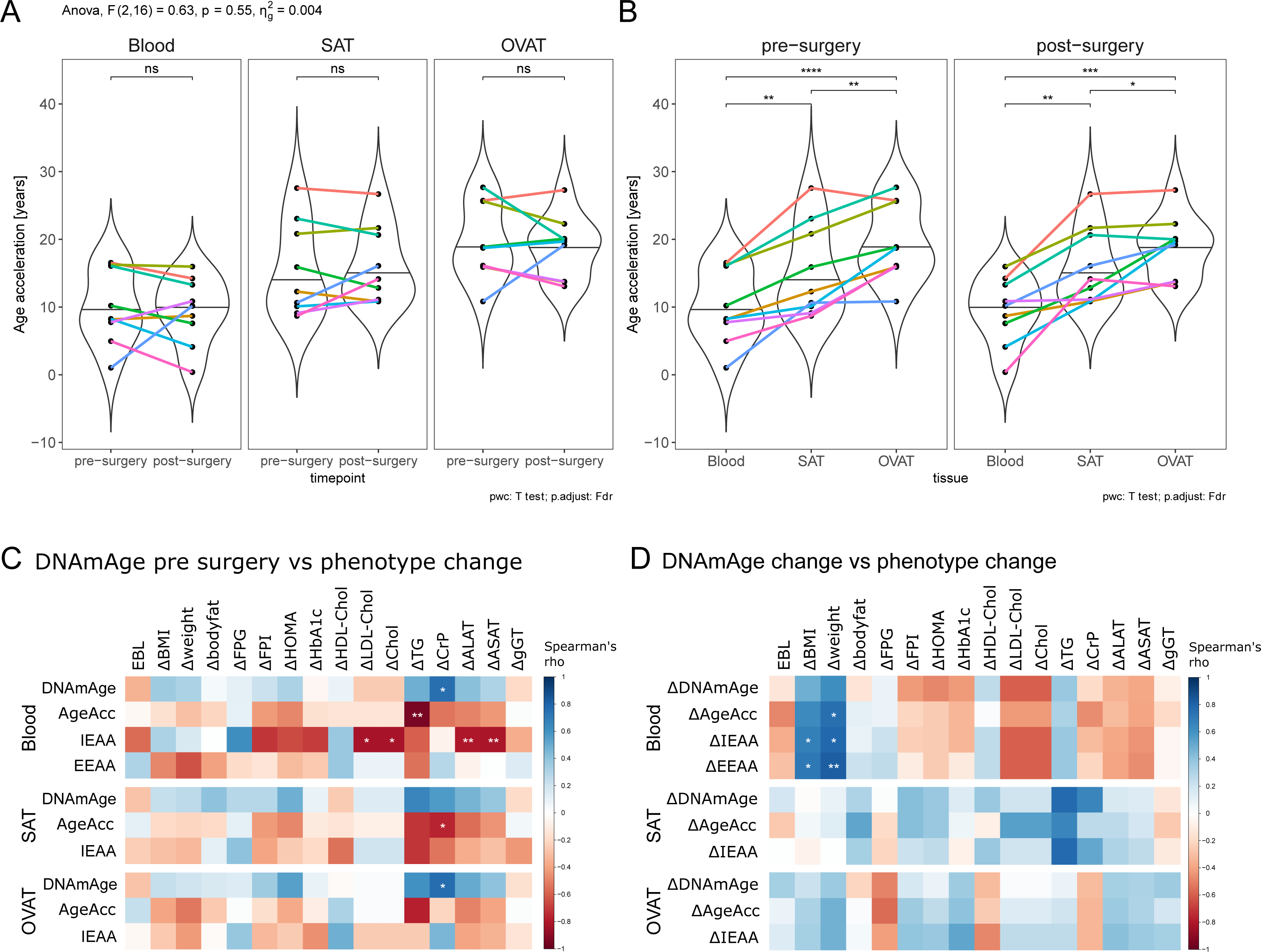
DNA methylation age after surgery and across tissues. **A/B)** Violin plots compare age acceleration (ageAcc) for blood, subcutaneous adipose tissue (SAT) and omental visceral adipose tissue (OVAT) before and after surgery. Each individual is defined by one colour. With two-way ANOVA the effect of surgery and tissue type on ageAcc was tested, followed by pairwise paired Student’s t-test. Significance level was adjusted for multiple comparison by false discovery rate (FDR); ***FDR < 0.001, **FDR < 0.01, *FDR < 0.05, ns – not significant (FDR ≥ 0.05). **C)** Correlation matrix shows Spearman correlation between DNA methylation age (DNAmAge) measurements before bariatric surgery with phenotypic changes after surgery (post-pre) **D)** Correlation matrix shows spearman correlation between changes of DNAmAge measurements with phenotypic changes after surgery (post-pre). The colour depicts the Spearman correlation coefficient (red - negative correlation, blue - positive correlation). Nominal significant correlations (not adjusted for multiple comparisons) are marked, \*\**P* < 0.01, \**P* < 0.05. ALAT - alanine aminotransferase, ASAT - aspartate aminotransferase, BMI - body mass index, CRP – C-reactive protein, EBL - excess BMI loss, EEAA - extrinsic epigenetic age acceleration, FPG - fasting plasma glucose, FPI - fasting plasma insulin, gGT - gamma glutamyltransferase, HDL - high density lipoprotein, HOMA - homeostasis model assessment IEAA - intrinsic epigenetic age acceleration, LDL - low density lipoprotein

### DNAmAge measures correlate with phenotype change after surgery

Besides age acceleration, other measures of DNAmAge are the intrinsic epigenetic age acceleration (IEAA) as the residual obtained after regressing chronological age and DNAmAge, and the extrinsic epigenetic age acceleration (EEAA) as the residuals obtained after regressing chronological age and DNAmAge adjusted for cell counts. We calculated Spearman’s rank correlation for the phenotype change (post – pre surgery) with DNAmAge measured before surgery (Figure 5C) as well as the change of DNAmAge measures (post-pre) (Figure 5D). We found a negative correlation of the change in triglyceride serum levels after surgery with the ageAcc in blood before surgery (R = -0.94, *P* < 0.01). Furthermore, IEAA in blood before surgery correlated negatively with the change of the liver enzyme serum levels of ALAT (R = -0.8, *P <* 0.01) and ASAT (R = -0.82, *P <* 0.01) after surgery. There were further correlations of the change in LDL-cholesterol and total cholesterol with IEAA pre-surgery in blood, and the change of CRP correlated positively with DNAmAge in blood and OVAT and negatively with age acceleration in SAT before surgery (all *P <* 0.05). The change of DNAmAge measures correlated in blood with weight and BMI loss. Best correlation was found with the change of EEAA and weight loss after surgery (R = 0.82, *P <* 0.01). However, correlations did not withstand correcting for multiple testing by FDR.

## 4. Discussion

The aim of our study was to uncover methylation changes in blood reflecting metabolic changes occurring in AT after bariatric surgery. We identified cross-tissue methylation patterns, which were validated in a separate set of individuals undergoing bariatric surgery. We further show that methylation changes are annotated to genes differentially expressed in AT after bariatric surgery. The major strength of our study is the pairing of blood and fat samples acquired both before and after surgery-induced weight loss, sourced from the same individuals, which reduces the effect of inter-individual variability.

We identified distinct patterns of DNA methylation changes after surgery both between blood and AT and between the two fat depots. In general, the quantity and the level of methylation changes was the highest in OVAT, followed by SAT and blood. Interestingly, hypomethylation prevailed in blood post-surgery, whereas AT showed more hypermethylated DMRs. This aligns with previous studies by Talukdar et al.^27^, who identified more hypomethylated blood DMPs/DMRs associated with surgery-induced weight loss. However, Benton et al.^26^, who investigated genome-wide DNA methylation in SAT and OVAT from women with obesity before and after RYGB, found more hypomethylated DMPs after surgery, which is contrary to our finding in AT. This discrepancy might be due to different factors, such as different array types, data analysis, and most importantly differences of patient phenotypes across different cohorts. For example, Benton et al. analysed the DNA methylation only in women, while we studied men and women. Moreover, Benton et al. analysed methylation on single CpG resolution only, while we concentrated on DMRs, potentially suggesting a stronger functional consequence. Nevertheless, our study suggests that adipose tissue, particularly OVAT, is more susceptible to epigenetic remodelling after bariatric surgery compared to blood.

As expected, we predominantly observed tissue-specific methylation changes after surgery with more than 50% of the DMRs after surgery showing no overlap across all three tissues. The annotated genes of the most significant tissue-specific DMRs were enriched for GO terms specific for the respective tissue, such as hemopoiesis and leukocyte activation/differentiation for blood. This supports the finding of Huang et al.^29^ in healthy individuals who found that discordantly methylated genes were enriched for biological processes related to immune cell activation, lymphocyte differentiation, and blood coagulation. Interestingly, although distinct methylation changes were observed for SAT and OVAT, both were annotated to genes enriched for similar biological processes, such as developmental process and morphogenesis. This indicates that bariatric surgery affects AT signature and biological function changes independently of the fat depot.

Overlapping DMRs between blood and AT are related to immune system processes independent of their effect direction. More than half of the cross-tissue DMRs showed methylation changes with discordant effect directions between blood and both ATs. Enriched GO terms for the genes associated with these DMRs included hemopoiesis and cytokine production. This could potentially signify a reduction of immune cell presence in AT following bariatric surgery, mirroring a cellular remodelling of the AT after surgery^70^. Also Talukdar et al.^27^ found in their epigenome-wide profiling of blood DNA methylation an enrichment of weight loss associated blood DMPs in immune and inflammatory signalling pathways. Furthermore, a decreased systemic inflammatory state after surgery^10^ is supported by the significantly reduced CRP levels in our cohort. Interestingly, the GO term immune system process was also enriched for the concordant DMR related genes, indicating immune processes that are similarly affected in both blood and AT after surgery.

Most identified cross-tissue DMRs are located in regulatory regions, indicating that surgery-induced methylation changes lead to transcriptional changes in all analysed tissues. In fact, we identified 14 cross-tissue DMRs, which were related to differentially expressed genes in at least one fat depot after surgery-induced weight loss. Amongst them, most DMRs are predicted to be within regulatory regions for gene transcription in both adipocytes and the blood cells. However, of those, the DMR at *PIK3AP1* was the only one related to differential gene expression in both SAT and OVAT. The observed downregulation of *PIK3AP1* mRNA expression in both depots after weight loss was in line with the observed hypermethylation of the promoter region in AT and was additionally mirrored in blood. Through regulation of Phosphoinositide 3-Kinase (PI3K) activity, PIK3AP1 influences diverse cellular functions. It plays a role in immune cell signalling by acting as an adaptor molecule in the signal transduction pathways of B-cells^71,72^ and studies have suggested associations of PIK3AP1 with inflammatory processes^73^. Moreover, in line with our initial findings, two independent studies found *PIK3AP1* gene expression was continuously downregulated in SAT of women post bariatric surgery^15,62^ suggesting a potential role in AT function and metabolic regulation. Further supporting this, *PIK3AP1* gene expression in SAT correlated with obesity related traits. While requiring further validation, *PIK3AP1* might be a promising candidate gene in blood reflecting AT function.

Previous studies have shown an attenuated epigenetic age in blood after bariatric surgery and lifestyle induced weight loss^27,34,36^. Additionally, associations between DNAmAge in liver^31^ or the visceral AT^32^ and BMI have been noted. However, in our study, the average distance from chronological age (ageAcc) remained unchanged after surgery in all tissues. This may be attributed to power limitations given by our relatively small sample size. Moreover, the age range in our cohort between 26 and 64 years might also contribute to this observation. Nevalainen et al.^74^, who examined the relation between BMI and accelerated epigenetic aging in blood samples of three different age groups, found that the association between BMI and accelerated epigenetic aging in blood was evident only in the middle-aged group (40-49 years), not in younger adults (15–24 years) or those in the nonagenarian group (90 years). Thus, the effects of bariatric surgery on epigenetic age might vary based on the individual’s age. Although we did not observe a significant difference in ageAcc between pre- and post-surgery, our findings indicate that greater weight loss was linked to a stronger reduction in ageAcc in blood.

Moreover, we found ageAcc to be consistently increased in AT compared to blood samples, suggesting a higher biological age of AT compared to the constantly renewing blood cells. In general, we observed significant correlations of ageAcc between tissues. However, while the correlation between ageAcc of SAT and OVAT remained unchanged post-surgery, we found a stronger correlation between blood ageAcc and AT ageAcc before surgery than after surgery. Huang et al. also observed that the DNA methylation profile of AT in individuals with higher BMI becomes more similar to that of blood^29^. Thus, we hypothesise that AT in individuals with a higher BMI (before surgery) might exhibit increased infiltration of immune cells, leading to a more similar epigenetic pattern between blood and AT in contrast to the state after surgery, when both BMI and inflammation are reduced.

The major limitation of our study was the small sample size attributed to the challenges associated with collecting samples from repeated surgeries. Another limitation arising from the need for a two-step bariatric surgery is that we observe partially marginal weight loss (EBL < 50%) in our cohort. As a result, we cannot distinguish whether the observed methylation and gene expression differences are only induced by weight loss or influenced by other factors resulting from the surgery. Additionally, our analysis involved whole tissue samples with varying cell composition. While we did not find differences in the blood cell counts after surgery, we cannot say that this hold true for adipose tissue samples, making it difficult to discern if methylation differences are due to varying cell compositions.

In the future, it might be interesting to explore methylation changes in blood derived from adipose tissue, especially through cell free DNA or extracellular vesicles, providing a new non-invasive tool to monitor adipose tissue health and dysfunction.

## Conclusion

After analysing the methylation changes induced by bariatric surgery in blood and AT, we identified cross-tissue DMRs related to the immune system and observed a higher epigenetic age acceleration in AT compared to blood independent of bariatric surgery. Our findings suggest that blood methylation patterns might mirror the inflammatory state of AT. Further investigation is needed to examine whether blood methylation of identified genes such as *PIK3AP1* could serve as reliable biomarker for the inflammatory status of AT.

## Data Availability

All data produced in the present study are available upon reasonable request to the authors.

## Abbreviations

ageAcc: age acceleration
ALAT: alanine aminotransferase
AIM2: absent in melanoma 2
ASAT: aspartate aminotransferase
AT: adipose tissue
BMI: body mass index
CRP: C-reactive protein
DEG: differentially expressed gene
DMP: differentially methylated position
DMR: differentially methylated region
DNAmAge: DNA methylation age
EBL: excess BMI loss
EEAA: extrinsic epigenetic age acceleration
FPG: fasting plasma glucose
FPI: fasting plasma insulin
gGT: gamma glutamyltransferase
GO: gene ontology
HDL: high density lipoprotein
HOMA: homeostasis model assessment
IEAA: intrinsic epigenetic age acceleration
LDL: low density lipoprotein
OVAT: omental visceral adipose tissue
NXN: nucleoredoxin
PCA: principal component analysis
PI3K: phosphoinositide 3-kinases
PIK3AP1: phosphoinositide 3-kinase adapter protein 1
RYGB: Roux-en-Y gastric bypass
SAT: subcutaneous adipose tissue
T2DM: Type II diabetes mellitus

## Author contributions

MK, AH and L Müller initiated, conceived, and designed the study. L Müller, AH, PK and MK wrote the first manuscript draft. AH, TH, L Müller and SB performed statistical and bioinformatics analysis. MB, RC, AD conducted the LOBB and collected samples and phenotypes. AG, WS, HD, FN, CW performed RNA sequencing of adipose tissue samples. JZ and L Massier conducted meta-analysis of *PIK3AP1* gene expression associations with clinical traits. AH, PK, MK, RC, MS, and MB supported the critical data interpretation and reviewed the manuscript. All authors read and approved the final manuscript.

## Declaration of competing interest

MB received honoraria as a consultant and speaker from Amgen, AstraZeneca, Bayer, Boehringer-Ingelheim, Lilly, Novo Nordisk, Novartis, and Sanofi. All other authors declare no conflicts of interest.

## Acknowledgements

We thank all patients and their families for participating in this study. Moreover we acknowledge excellent technical assistance by Ines Müller.

The work is supported by the DFG (German Research Foundation) – Projektnummer 209933838 – SFB 1052 (project B1, B3) and by Deutsches Zentrum für Diabetesforschung (DZD, Grant: 82DZD00602). The German Diabetes Center is funded by the German Federal Ministry of Health (Berlin, Germany) and the Ministry of Culture and Science of the state North Rhine-Westphalia (Düsseldorf, Germany) and receives additional funding by the German Federal Ministry of Education and Research (BMBF) through the German Center for Diabetes Research (DZD e.V.). The funders had no role in the design of the study; in the collection, analyses, or interpretation of data; in the writing of the manuscript; or in the decision to publish the results.

## Notes

### Author Declarations

The study was approved by the Ethics Committee of the University of Leipzig (approval no: 159-12-21052012, 017ek-2012) and performed in accordance with the Declaration of Helsinki.

