## Supplemental Figures for "Blood methylation pattern reflects epigenetic remodelling in adipose tissue after bariatric surgery"

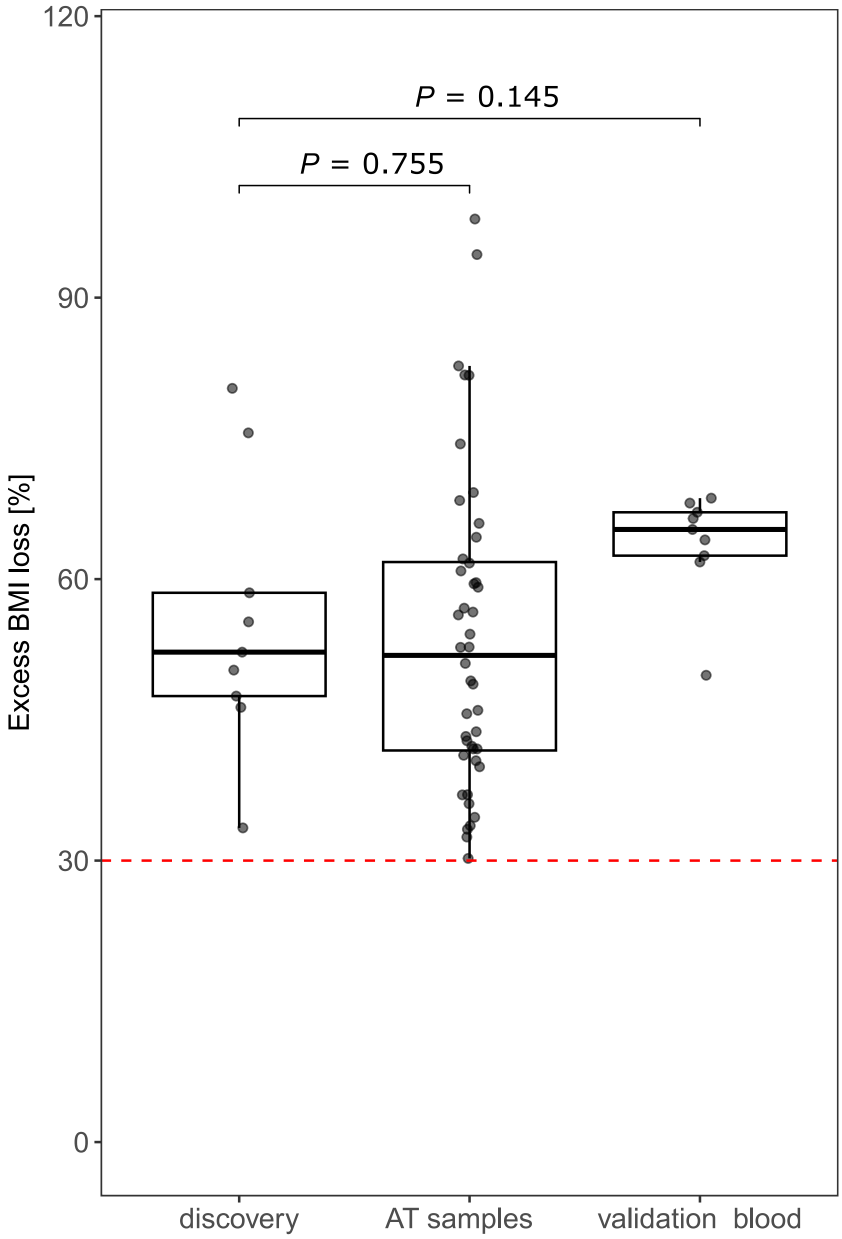


**Figure S1|** Boxplot shows comparison of excess BMI loss (EBL, %) between discovery sample set and validation sample sets. *P* represents significance level of unpaired Student's t-test.

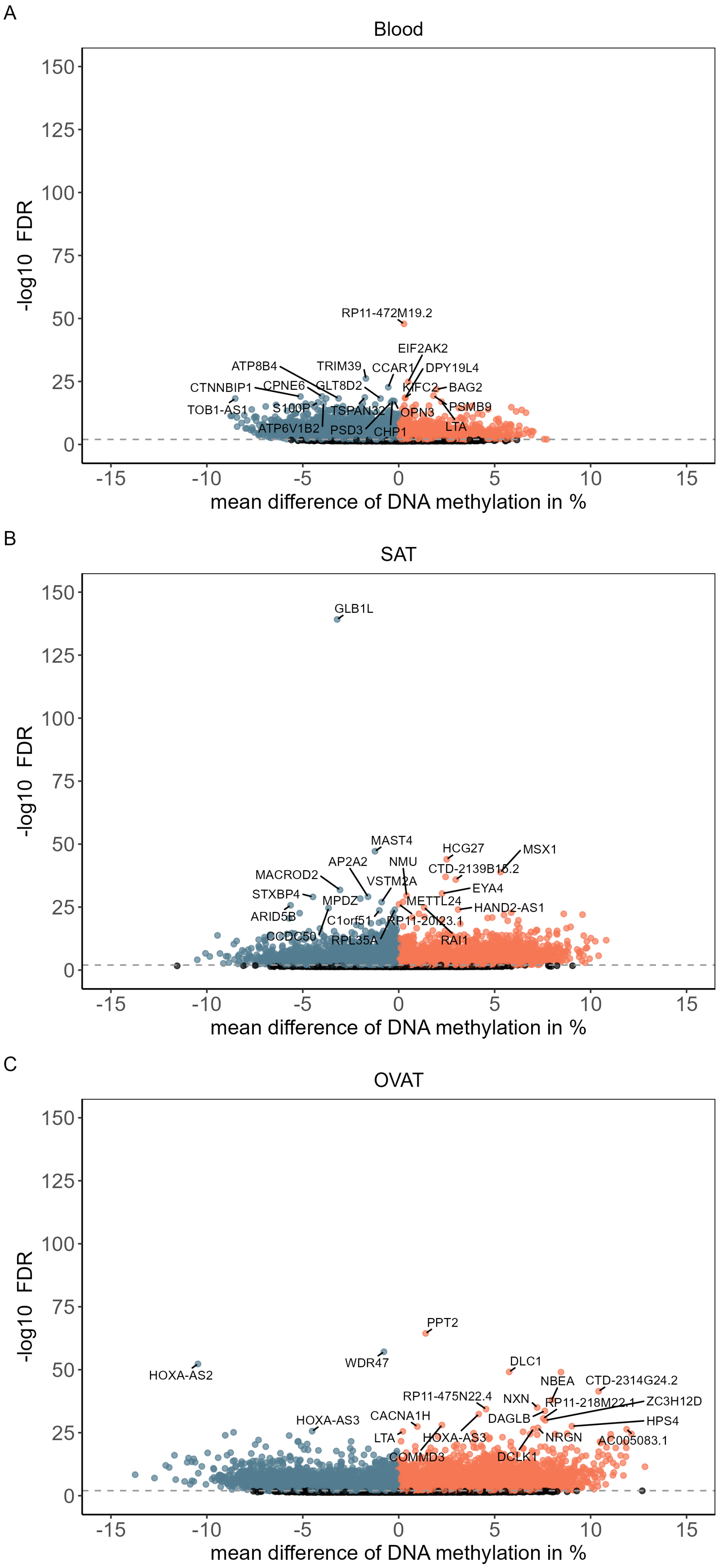


**Figure S2|** Volcano plots of DMRs in blood (A), subcutaneous adipose tissue (B), and omental visceral adipose tissue (C). Blue points represent hypomethylated DMRs (mean methylation difference < 0, FDR < 0.01) and red points represent hypermethylated DMRs (mean methylation difference > 0, FDR < 0.01) after surgery-induced weight loss. Black points show DMRs with FDR between 0.01 and 0.05. Top 20 DMRs (ranked by FDR) are labelled with the annotated gene.

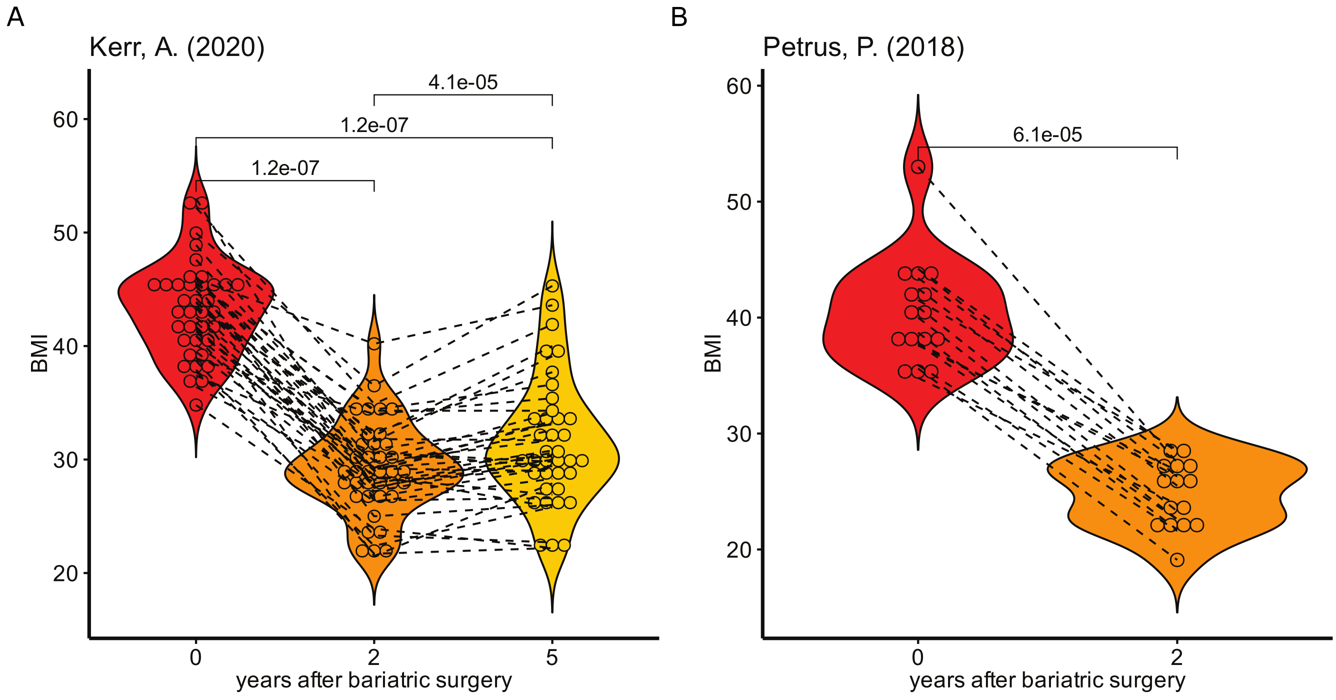


**Figure S3|** Violin Plots indicate weight loss after bariatric surgery in both studies of A) Kerr et al. (2020) and B) Petrus et al. (2018).

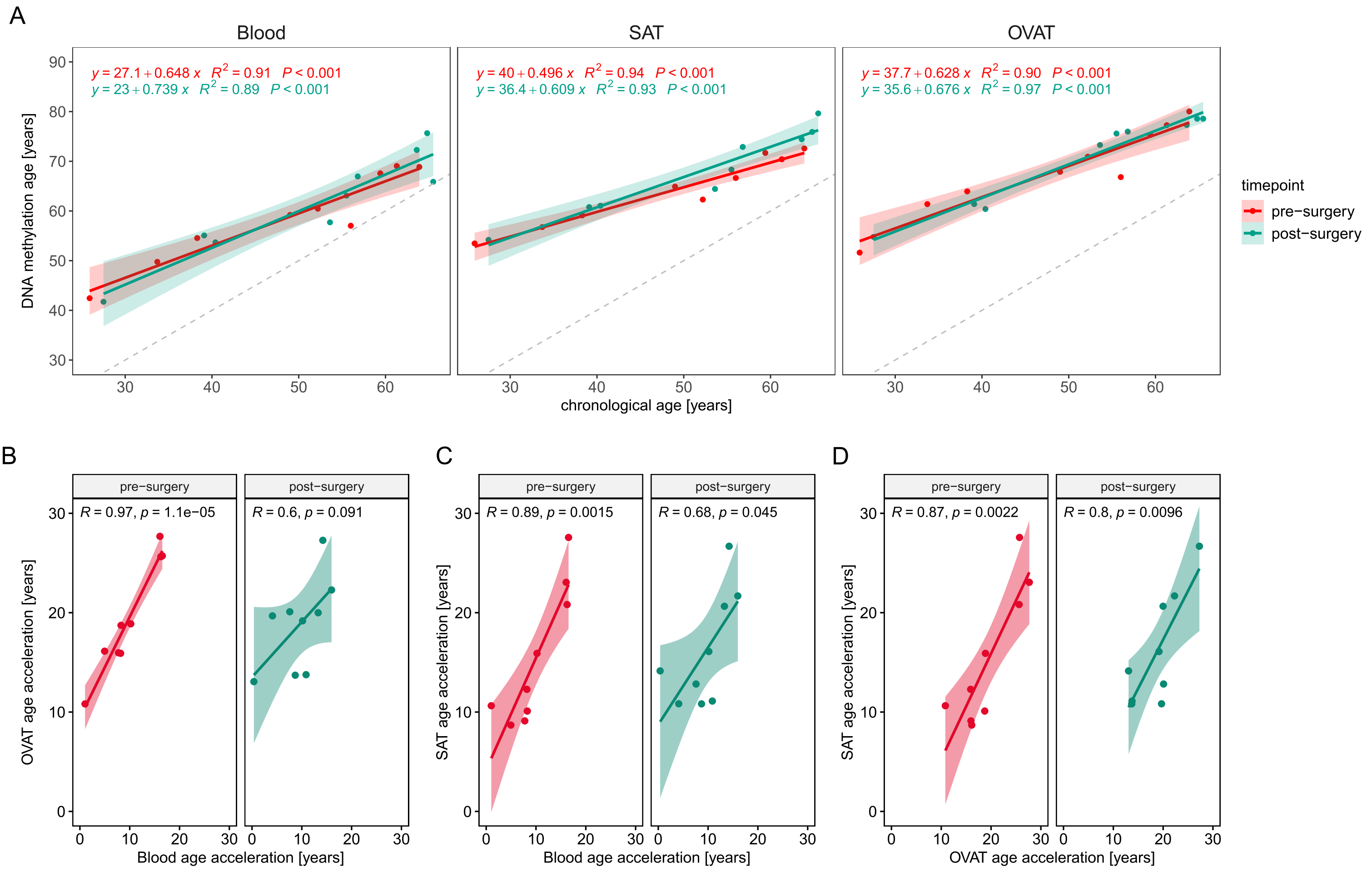


**Figure S4|** A) Linear regression of DNA methylation age (DNAmAge) ~ chronological age for blood, subcutaneous adipose tissue (SAT) and omental visceral adipose tissue (OVAT). Formula of regression line, fit ($R^2$) and significance level (*P*) are depicted for the associations pre-surgery in red and post-surgery in green. A grey dashed line visualises where DNAmAge is equivalent to the chronological age. B-D) Scatter plots show correlation of age acceleration (ageAcc) between tissues for pre and post-surgery. Spearman correlation coefficient (R = rho) and unadjusted *P* of correlation are depicted. A linear regression line is added. Correlations were calculated for ageAcc between B) blood and OVAT, C) blood and SAT, and D) SAT and OVAT.

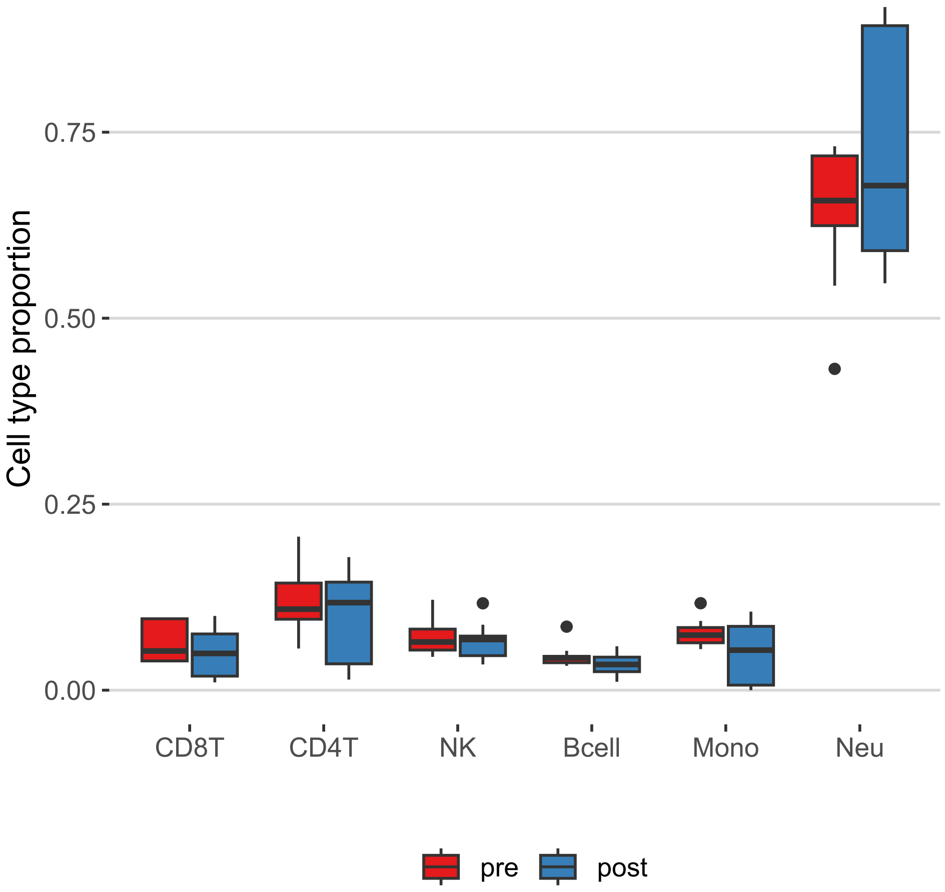


**Figure S5|** Boxplot shows proportions of cell type populations (CD8T, CD4T, natural killer cells (NK), B-cells, monocytes, and neutrophils) between pre- and post-surgery samples.

A

| 1 (22) | 2 (9) | 3 (9) | 4 (8) | 5 (7) | |
|---|---|---|---|---|---|
| 0.6 | 0.004 | 0.9 | 0.8 | 1 | Sample_Well |
| 0.03 | 6e−06 | 8e−07 | 3e−06 | 1e−09 | Patient |
| 0.5 | 0.9 | 1 | 0.9 | 0.9 | Group |
| 0.3 | 0.8 | 0.01 | 0.9 | 0.08 | sex |
| 0.6 | 0.6 | 0.4 | 0.03 | 0.02 | Age |
| 0.6 | 0.1 | 0.3 | 0.3 | 1 | weight |
| 0.4 | 0.3 | 0.04 | 0.2 | 0.03 | height |
| 0.7 | 0.2 | 0.6 | 0.6 | 0.8 | BMI |
| 0.3 | 0.9 | 0.07 | 0.9 | 0.5 | BMIgroup |
| 0.2 | 0.7 | 0.1 | 0.4 | 0.01 | bodyFat |
| 0.8 | 0.6 | 1 | 0.4 | 0.2 | DiabetesState |
| 0.6 | 0.004 | 0.9 | 0.8 | 1 | Array |
| 0.005 | 0.8 | 0.7 | 1 | 0.8 | Slide |
| 7e−09 | 0.9 | 0.6 | 0.8 | 0.6 | CD8T |
| 4e−08 | 0.5 | 0.5 | 0.9 | 0.9 | CD4T |
| 0.1 | 0.8 | 0.03 | 0.7 | 0.2 | NK |
| 1e−04 | 0.7 | 0.2 | 0.5 | 0.9 | Bcell |
| 3e−04 | 0.9 | 0.09 | 0.6 | 0.7 | Mono |
| 2e−08 | 0.6 | 0.3 | 1 | 0.9 | Neu |

Principal Components (Variation)

B

| 1 (29) | 2 (14) | 3 (10) | 4 (6) | 5 (5) | |
|---|---|---|---|---|---|
| 0.02 | 0.3 | 0.02 | 0.6 | 0.5 | Sample_Well |
| 0.1 | 0.01 | 0.008 | 0.001 | 1e−05 | Patient |
| 0.7 | 0.2 | 0.5 | 0.7 | 0.6 | Group |
| 0.1 | 0.8 | 0.01 | 0.005 | 0.9 | sex |
| 0.3 | 0.05 | 0.05 | 0.5 | 0.1 | Age |
| 0.9 | 0.9 | 0.4 | 0.6 | 0.5 | weight |
| 0.3 | 0.6 | 0.07 | 0.07 | 0.7 | height |
| 0.8 | 0.8 | 0.5 | 0.3 | 0.5 | BMI |
| 0.01 | 0.4 | 0.2 | 0.4 | 0.4 | BMIgroup |
| 0.4 | 0.9 | 0.01 | 0.3 | 0.5 | bodyFat |
| 1 | 0.1 | 0.2 | 0.6 | 0.8 | DiabetesState |
| 0.02 | 0.3 | 0.02 | 0.6 | 0.5 | Array |
| 0.7 | 0.8 | 0.7 | 0.8 | 0.2 | Slide |

Principal Components (Variation)

C

| 1 (27) | 2 (20) | 3 (7) | 4 (5) | 5 (5) | |
|---|---|---|---|---|---|
| 0.2 | 0.7 | 0.4 | 0.4 | 0.2 | Sample_Well |
| 1 | 0.2 | 0.02 | 0.04 | 0.003 | Patient |
| 0.005 | 0.6 | 0.9 | 0.5 | 0.4 | Group |
| 0.9 | 0.5 | 0.001 | 0.6 | 0.04 | sex |
| 0.4 | 0.2 | 0.1 | 0.2 | 0.4 | Age |
| 7e−05 | 0.7 | 0.8 | 0.5 | 0.6 | weight |
| 0.7 | 0.4 | 0.06 | 0.9 | 0.06 | height |
| 2e−04 | 0.8 | 0.9 | 0.9 | 0.8 | BMI |
| 0.06 | 0.8 | 0.09 | 0.9 | 0.5 | BMIgroup |
| 0.2 | 0.8 | 0.07 | 0.7 | 0.008 | bodyFat |
| 0.7 | 0.009 | 0.6 | 0.6 | 0.3 | DiabetesState |
| 0.2 | 0.7 | 0.4 | 0.4 | 0.2 | Array |
| 0.4 | 0.8 | 0.9 | 0.7 | 0.2 | Slide |

Principal Components (Variation)

**Figure S6|** Heatmaps show principal component analyses (PCA) of normalised EPIC array data in blood (A), subcutaneous adipose tissue (B), and omental visceral adipose tissue (C).

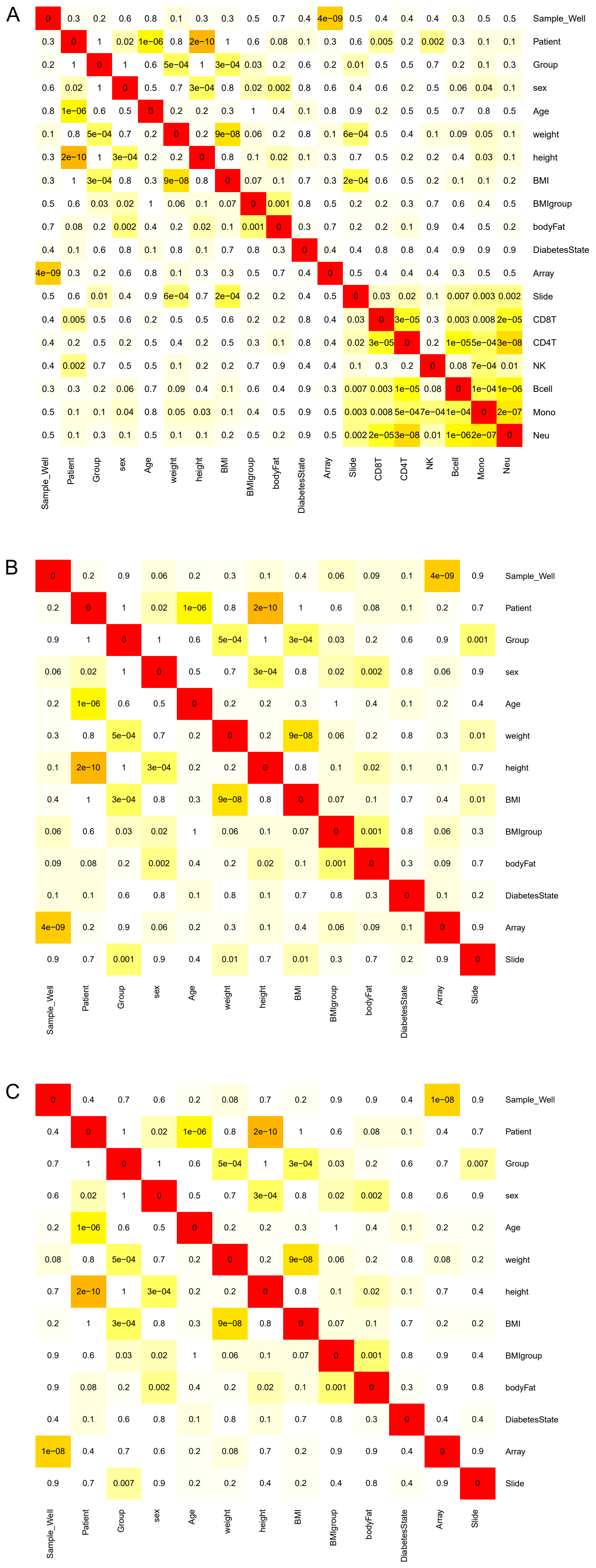


**Figure S7|** Correlation matrix to identify possible confounders in blood (A), subcutaneous adipose tissue (B), and omental visceral adipose tissue (C).

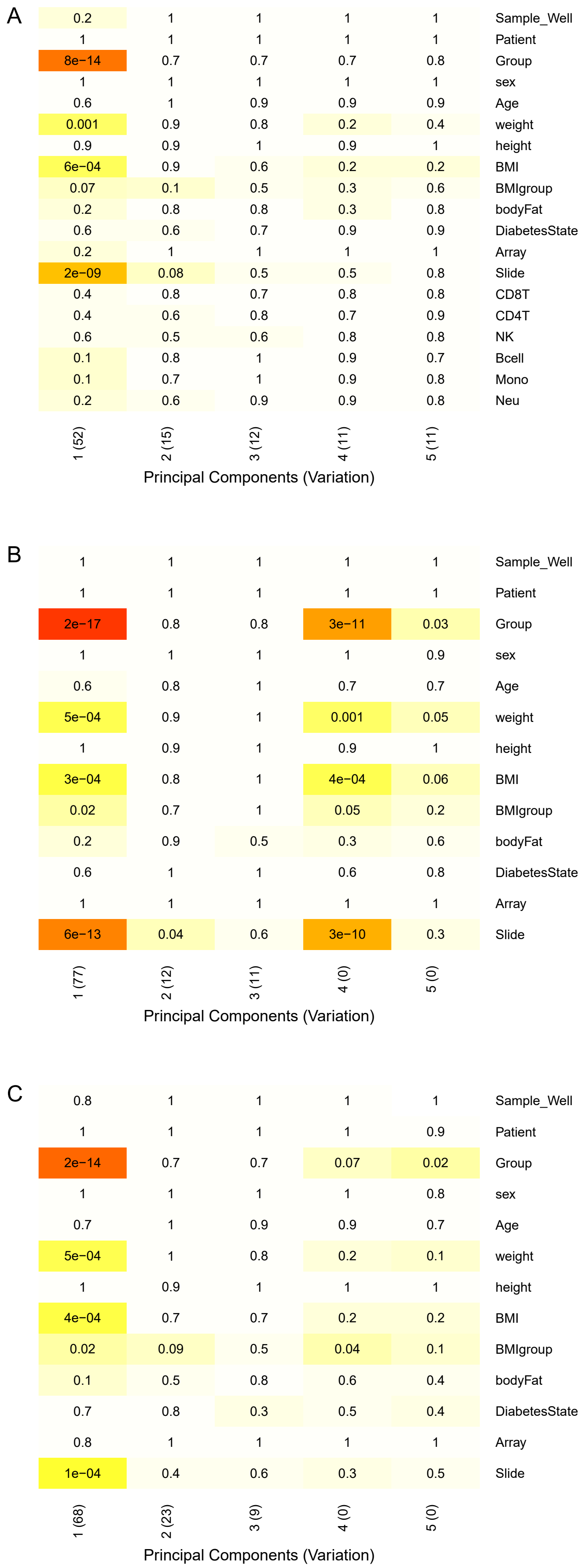


**Figure S8|** Heatmaps show principal component analyses of EPIC array data adjusted for array and patient in blood (A), subcutaneous adipose tissue (B), and omental visceral adipose tissue (C).

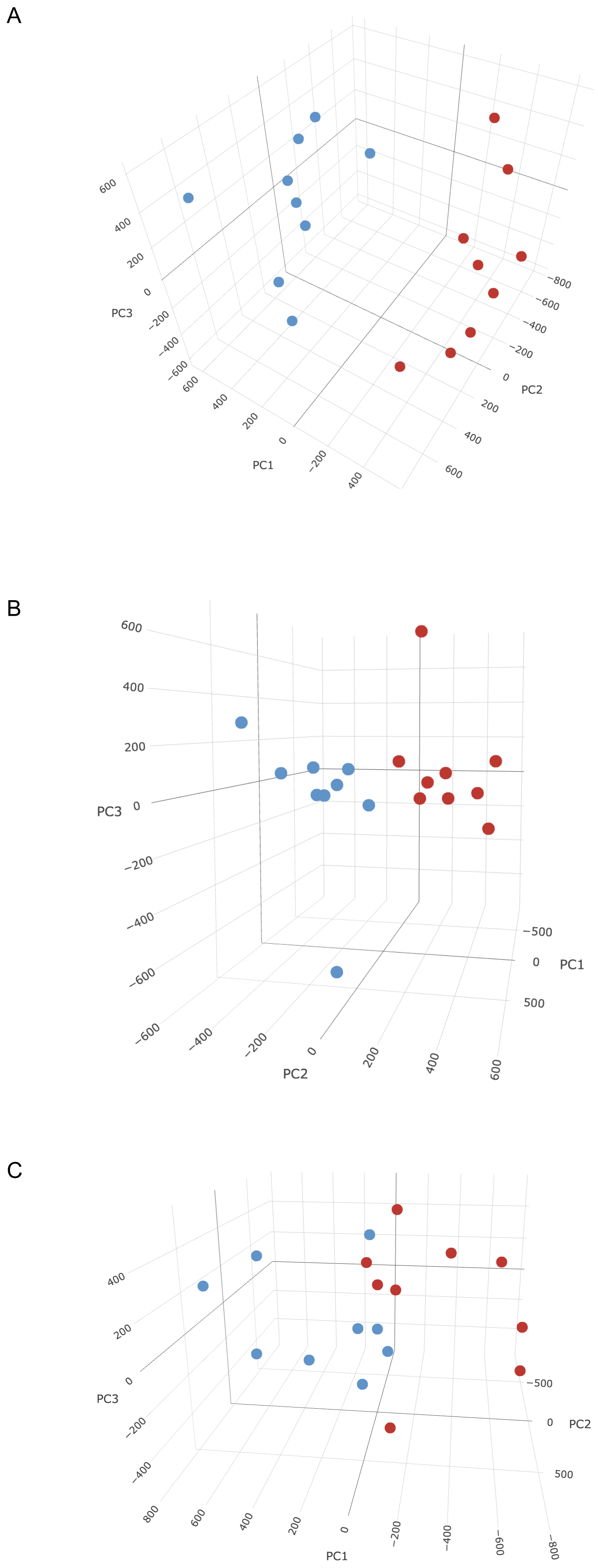


**Figure S9|** PCA plot in blood (A), subcutaneous adipose tissue (B), and omental visceral adipose tissue (C).
