## Supplemental Information for "Blood methylation pattern reflects epigenetic remodelling in adipose tissue after bariatric surgery"

**Supplementary information**

*1. Phenotyping of Leipzig Obesity BioBank (LOBB)*

Phenotyping of the LOBB includes assessment of age, sex, BMI (kg/m²), body fat mass (%) detection using bioelectrical impedance analysis, and metabolic biochemical assessment such as fasting plasma glucose (FPG, mmol/l) and fasting plasma insulin (FPI, pmol/l), HbA1c levels (%), triglycerides (TG, mmol/l), high- and low-density lipoprotein (HDL/LDL, mmol/l) cholesterol, liver enzymes alanine and aspartate aminotransferase (ALAT/ASAT, mmol/l) and gamma-glutamyl transferase (gGT, mmol/l), as well as C-reactive protein levels (CRP, µg/ml). Homeostatic model assessment for insulin resistance (HOMA-IR) was calculated as described elsewhere^1^.

*2. Sample collection and nucleic acid isolation*

AT samples (SAT and OVAT) were taken during elective laparoscopic abdominal surgery as previously described^2,3^. EDTA blood samples were collected prior to each metabolic surgery. For the validation analysis, the second blood sample was collected at the follow-up visit 24 months post-surgery. All samples were immediately frozen and stored at -80 °C until further processing.

For DNA isolation from EDTA blood and AT samples, the DNeasy blood and tissue kit (Qiagen, Hilden, Germany) was used following manufacturer's instructions. Yield, integrity and quality of DNA samples were assessed using Quantus™ (Promega, Wisconsin, USA) and Nanodrop 2000 spectrometer (Thermo Fisher Scientific, MA, USA). Total RNA was extracted from SAT and OVAT using RNeasy Lipid tissue Mini Kit (Qiagen, Hilden, Germany) as described before^4^.

*3. Genome-wide DNA methylation data processing*

Probes that did not surpass a detection *P*-value > 0.01 in more than 1% of all samples, cross-reactive probes detected using the *maxprobes* R package v0.0.2^5^, probes containing known single nucleotide polymorphisms (SNPs) at the CpG site, and probes located on chromosomes X and Y were excluded from further analysis. After probe exclusions, a total of 866,238 probes for the discovery sample set and 866,091 for the validation blood samples were retained and utilised in downstream statistical analyses.

For blood samples, cell type heterogeneity using the Houseman approach^6,7^, adapted to EPIC arrays by Salas et al.^8^ with the application of the *FlowSorted.Blood.EPIC^9^* R package v2.4.2 was assessed. None of the cell type populations (CD8T, CD4T, natural killer cells (NK), B-cells, monocytes, and neutrophils) exhibited significant differences between pre- and post-surgery samples (Suppl. Fig. 5). Batch effects and/or technical sources of variation (array, well and slide) were monitored by assessing the association between the top principal components and possible batches using the R package *swamp* v1.5.1^10^ (Suppl. Fig. 6A-C). The technical parameters array and sample-well demonstrated a significant batch effect in most cases. Since both technical batches were strongly interrelated (Suppl. Fig. 7A-C) we adjusted for the array-batch effect. Statistically significant associations were observed between the patient (paired samples) batch. Accounting for the patient-batch reduces noise in the data by accounting for individual variance. Subsequently, the data were corrected for the array and patient batches using the R package *limma* v3.56.1^11^. To ensure that batch corrections were not biased by tissues, the analyses were performed independently for each tissue. After batch correction, there was no significant impact on the variance of the data for the cell types in the blood (Suppl. Fig. 8A-C), thus no additional correction was necessary in this regard. PCA separation of pre- and post-surgery clusters in blood, SAT and OVAT was noted after adjustment (Suppl. Figure 9A-C).

4. RNA sequencing analyses

Raw sequencing reads were pre-processed using *fastp* (v0.20.0)^12^ with a minimum read length of 18 nts and a quality cut-off of 20. To align the reads to the reference genome (assembly GRCh38.p13, GENCODE release 32^13^) and quantify gene-level expression, we utilized the pseudoaligner *kallisto^14^*. In cases, where samples had more than 20 million read counts, downsampling was performed to achieve a consistent count of 20 million reads using the R package *ezRun* (v3.14.1; <https://github.com/uzh/ezRun>, accessed on 23 March 2022).

*6. Methylation age analysis*

DNA methylation age (DNAmAge) was calculated using the R package *methylclock* v.1.6.0^15^ For DNAmAge calculation normalised (not batch corrected) beta values from the EPIC array data of 353 CpGs (Horvath’s clock^16^) were used, since calculation includes a special cell count adjustment method. In blood all CpG methylation sites necessary to estimate DNAmAge were covered with our methylation data. For SAT and OVAT, 19 CpGs were not covered by EPIC array, but it was already shown to not affect DNAmAge prediction^17^. DNAmAge acceleration (ageAcc) is calculated as the difference of chronological age given by the date of birth and DNAmAge (ageAcc = DNAmAge - chronological age).

*7. Correlation with clinical traits*

In essence, data of previously published microarray and RNA sequencing was reanalysed using established pipelines: DESeq2 v1.34^18^ for RNA sequencing and either oligo v1.58.0^19^ or lumi v2.24.0 ^20^ (depending on the array type) in conjunction with limma v3.50.3^11^ in R for microarray analysis. Spearman correlation was computed using the rcorr function within Hmisc v5.0-1^21^, while meta-analysis was executed and visualized through the metacor and forest.meta functions within the meta v6.1-1^22^ package. To compare values before and after weight loss, normalized and log2-transformed data were compared applying a Wilcoxon signed rank test for paired samples.
